# Analysis of overdispersion in airborne transmission of Covid-19

**DOI:** 10.1101/2021.09.28.21263801

**Authors:** Swetaprovo Chaudhuri, Prasad Kasibhatla, Arnab Mukherjee, William Pan, Glenn Morrison, Sharmistha Mishra, Vijaya Kumar Murty

**Affiliations:** Institute for Aerospace Studies, University of Toronto; Nicholas School of Environment, Duke University; Global Institute of Health, Duke University; Gillings School of Global Public Health, University of North Carolina; Department of Medicine, Division of Infectious Diseases, University of Toronto; The Fields Institute for Research in Mathematical Sciences; Department of Mathematics, University of Toronto

## Abstract

Superspreading events and overdispersion are hallmarks of the Covid-19 pandemic. To gain insight into the nature and controlling factors of these superspreading events and heterogeneity in transmission, we conducted mechanistic modeling of SARS-CoV-2 transmission by infectious aerosols using real-world occupancy data from a large number of full-service restaurants in ten US metropolises. Including a large number of factors that influence disease transmission in these settings, we demonstrate the emergence of a stretched tail in the probability density function of secondary infection numbers indicating strong heterogeneity in individual infectivity. Derived analytical results further demonstrate that variability in viral loads and variability in occupancy, together, lead to overdispersion in the number of secondary infections arising from individual index cases. Our analysis, connecting mechanistic understanding of SARS-CoV-2 transmission by aerosols with observed large-scale epidemiological characteristics of Covid-19 outbreaks, adds an important dimension to the mounting body of evidence with regards to the determinants of airborne transmission of SARS-CoV-2 by aerosols in indoor settings.

## 1 Introduction

Superspreading events and overdispersion are now well-established characteristics of the Covid-19 pandemic, similar to SARS and other outbreaks of respiratory viruses [1]. The documented outbreaks at the Skagit Valley Chorale [2, 3], at a restaurant in Guangzhou [4, 5] and at a call center in Korea [6] are examples of superspreading events where one infectious index case led to tens of individuals infected within a few hours, at an order of magnitude higher than the basic reproduction number 2 ≤ *ℛ*_0_ ≤ 3.6 for the original SARS-CoV-2 variant [7]. The term “superspreading” has generally been ascribed to any event (index case and exposures) that led to more than the average number of secondary transmissions, and thus in probabilistic terms could refer to any number of secondary cases to the right of the expectation [1]. As such, it has been proposed that superspreading events are not exceptional events but an expected feature of an expected right-sided tail of the distribution of the basic reproduction number. When this right-hand tail is further skewed, with greater variability than expected leading to an uneven distribution, the term overdispersion is applied (statistical definition). In the context of communicable diseases, overdispersion refers to a non-random pattern of clustering, and which often include a large number of zero cases and a small number of larger outbreaks [8]. This pattern of overdispersion can be applied at the level of an event or index case (individual-level variation) [8], or in the context of networks (population-level via onward transmission chains) [9]: both fall under the broader epidemiological umbrella of heterogeneity. In the context of the former, data suggest that such superspreading events are characterized by overdispersion in SARS-CoV-2 transmissions, with 10-20% of index cases responsible for 80% of secondary cases [10].

Understanding the nature and characteristics of superspreading events is therefore key to understanding SARS-CoV-2 spread. Agent based modeling by Sneppen et al. [11] suggested that non-repeating, random contacts such as those in restaurants, bars etc. is a dominant contributor to SARS-CoV-2 spread. Chen et al. [12] argue that while social, and micro-environmental factors affect transmissibility, overdispersion could result from an intrinsic characteristic of certain viruses. The knowledge gap that remains is the extent to which each of these factors contribute to the observed distribution of secondary SARS-CoV-2 transmissions per index case, by connecting variability in each of the components with our mechanistic understanding of SARS-CoV-2 transmission.

Emerging data strongly suggest the importance of airborne transmission of SARS-CoV-2 by respiratory aerosols [13, 14, 15, 16, 17]. A large number of small respiratory droplets (when size *<* 100*µm* at point of exhalation) often referred as aerosols, as suggested by Prather et al. [18], can remain airborne in the liquid or semi-solid state [19], encapsulating the SARS-CoV-2 virus. As a result, the virus can remain infectious within the aerosols for substantial length of time [20]. Several studies have analyzed airborne transmission in specific micro-environments. Bourouiba et al. [21] identified that respiratory droplets and aerosols exhaled during violent expiratory events can travel long distances co-flowing with the moist air jet. Abkarian et al. [22], analyzed exhaled air flow during speech and how certain phonetics produce a train of puffs. Chen et al. [23] showed that for talking and coughing, short range airborne route dominates transmission of respiratory infections. Analyzing a respiratory droplet/aerosol laden cough jet, modeling by Chaudhuri et al. [19] showed that aerosols (droplet-nuclei) of initial size less than 50 *µ*m pose highest infection risk and variation in the corresponding viral load could lead to large variation in the number of secondary infections. Using well-mixed assumptions and the Wells-Riley model [24, 25], Buonanno et al. [26] proposed a quantitative risk assessment for specific micro-environments with asymptomatic infectious cases and suggested that instead of specific superspreaders it is a combination of several factors, including emission and exposure that lead to highly probable, superspreading events. Using similar well-mixed approach Bazant and Bush [27] proposed to restrict occupancy number and time spent in a room, to mitigate airborne transmission [27]. Schijven et al. estimated risk of infection resulting from sneezing, coughing, speaking, breathing, and singing [28] at different viral loads. Analysis by Bond et al. [29] quantified the importance of confinement in pathogen transport using “effective rebreathed air volume”. Further details on specific aspects of airborne disease transmission: including but not limited to aerosols, flow physics, respiratory droplet size distribution could be found in recent review and opinion articles [30, 31, 32, 33, 34]. Yet, to date mechanistic models describing airborne transmission have not been coupled with real-world distributions and occupancy information towards understanding the large scale features of disease dynamics, for e.g. overdispersion in transmissibility.

The overarching goal of this work is to utilize the mechanistic underpinnings of the airborne disease transmission to explore event-level overdispersion of SARS-Cov-2 spread, utilizing real-world inputs from a large number of social gatherings. The specific goals are:

1. Develop an algorithm based on aerosol dispersion with randomized inputs and available occupancy data to generate distribution of the number of secondary infections per infectious case.
2. Explore if observed patterns of overdispersion in secondary transmissions could be reproduced via simulations using the above algorithm.
3. Derive an analytical function (and not a fit) which can describe the probability density function of number of the secondary infections from the dynamics of the problem.
4. Identify the dominant variables that drive overdispersion and the resulting implications for mitigation measures.

To these ends, an aerosol spread model is solved over a hundred thousand random social-contact settings, utilizing real-world occupancy and area information coupled with realistic input distributions of viral-load and ventilation rates, to obtain the probability distributions of the number of secondary infections per infectious case, in those settings.

## 2 Method

In this section, first we describe the model to obtain the probability of infection resulting from inhalation of infectious aerosols generated from speaking and breathing, in an indoor, confined, and ventilated micro-environment. Next, we connect this model to an algorithm - a first of its kind to our knowledge, that accepts randomized inputs from distributions of viral load, exhaled aerosol size distribution, ventilation rate, speech and exposure time corresponding to specific inputs of number of people occupying specific indoor areas. The occupancy information is obtained from a large SafeGraph data-set of full-service restaurants from ten major US cities. The restaurants serve as our study setting for time-limited, point source transmission via mostly non-repeating, random contacts, in contrast to households or workplaces where repeated contacts are made with the same individuals and over longer periods of time. In this work, we focus on disease spread by asymptomatic infectious cases (asymptomatic at the time of disease spread therefore including presymptomatics) under the assumption that individuals with symptoms would not be engaged in indoor dining. Hence, we consider only speech and breath as the mechanisms by which respiratory aerosols are ejected into specific micro-environments. The algorithm developed for this work, is shown in Fig. 1.

**Figure 1:**
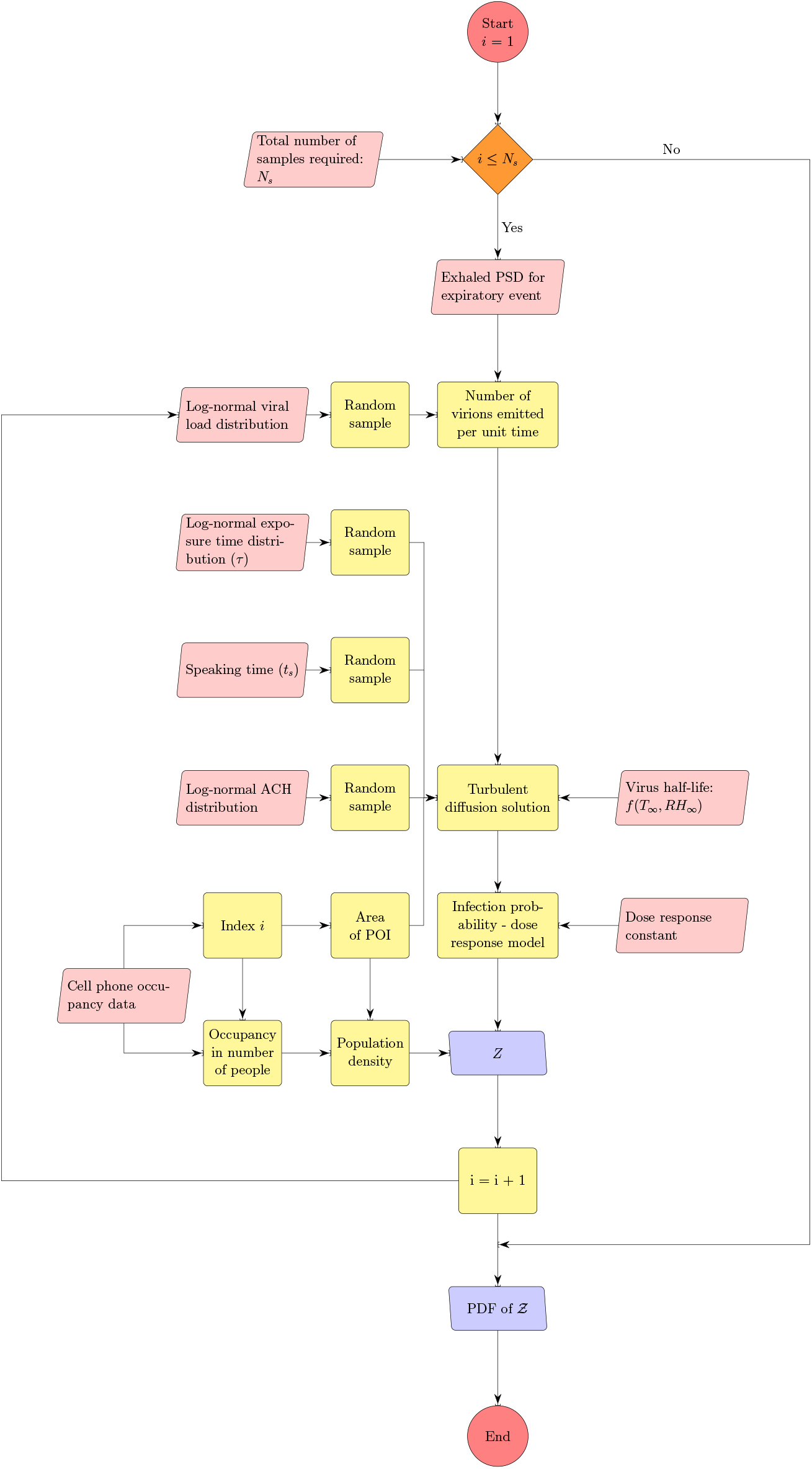
The algorithm for estimating distribution of secondary infections *Z*.

### 2.1 Aerosol dispersion from an infectious case

We use the standard, turbulent diffusivity based closure [35] to model the spatio-temporal evolution of ensemble averaged infectious aerosol concentration *c*_*a*_ (number of aerosol particles per unit volume of air associated with emissions from the infector), as shown by Eqn. 1. This approach was used by Drivas et al. [36] to model indoor concentration fields from point sources.

The turbulent diffusivity *D*_*T*_ in Eqn. 1 is a function of the air-change rate *a* and area of the space given by *A*. The second term on the R.H.S. is a sink term that accounts for removal of particles by ventilation (*a*: air change rate) and by deposition, while *w*_*d*_ is the deposition velocity and *V* being the volume of the indoor space.

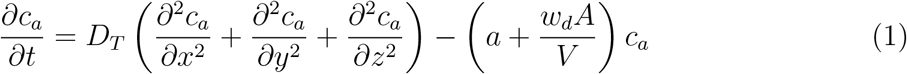

We treat the infectious case as a continuous point source. The initial condition is *c*_*a*_(*x, y, z*, 0) = 0 with reflection boundary conditions at the six walls. Since the virus remains embedded inside the infectious aerosols and since they are non-volatile, the ratio of the number of virions to the number of aerosol particles in any given volume of air can be assumed to remain constant post-ejection. To retain analytical tractability of the solution, in this work, we consider fast evaporation and a constant, post-evaporation, aerosol volume-averaged *w*_*d*_; see [23, 37, 19] for detailed evaporation and deposition considerations of respiratory droplets. Therefore, the ensemble averaged concentration of virus RNA copies per unit volume of air *c* is proportional to *c*_*a*_, or

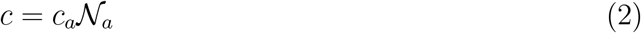

The average number of virions within an aerosol particle, *𝒩*_*a*_, is the constant of proportionality. Using the identity Eq. 2 we can immediately convert Eqn. 1 into an evolution equation for *c*, as shown in Eqn. 3:

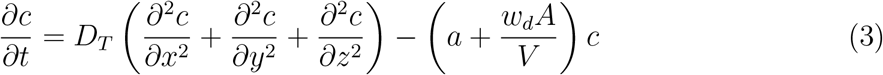

While one can use more complex approaches to model turbulent mixing and aerosol dispersion, the advantage of the relatively simple, yet sufficiently robust and accurate, diffusivity based closure is the analytical tractability and the inexpensive solution it offers. As will be seen later, such an attribute is pivotal to generating the large number of realizations of the *c* field, utilized in this work. As such, for a continuous point source 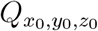 at (*x*_0_, *y*_0_, *z*_0_), the solution of the concentration field *c*(*x, y, z, t′*) at time *t′* was given by [36]:

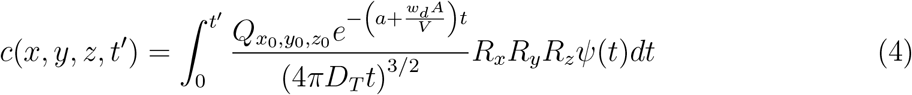

with wall reflection terms for *i, j, k* ≠ 0

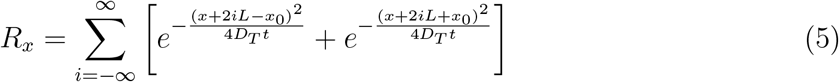

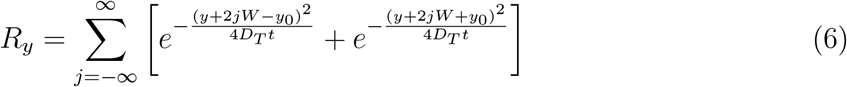

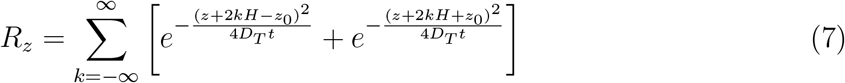

*L, W, H* are the length, width, and height of the room, respectively. Here, we have introduced a virus survivability function *ψ*(*t*) in the solution, as in Chaudhuri et al. [19]

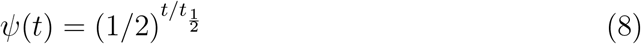

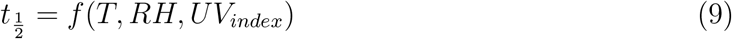

Here *T* is the temperature, *RH* is the relative-humidity and *UV*_*index*_ is the ultra-violet index inside the room of interest. Based on the experiments by Dabisch et al. [20] and calculator from DHS [38], for *T* = 21.7^*o*^*C, RH* = 0.50, *UV*_*index*_ = 0 i.e. typical ASHRAE recommended indoor air conditions, for SARS-CoV-2, half-life 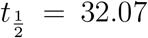 minutes. The SARS-CoV-2 virus half-life reduces monotonically with temperature and non-monotonically with relative humidity [39] like other enveloped viruses, as shown by Marr et al. [40].

The above solution given by Eqns. 4-7 (without *ψ*(*t*)) was validated by Cheng et al. [41] who released CO from a point source and measured its spatio-temporal dispersion characteristics inside typical built environments. Indeed aerosols deposit (accounted for in this paper) unlike CO, but since their motion is predominantly controlled by turbulent diffusion inside a room (turbulent diffusivity *D*_*T*_ *>>* molecular diffusivity of CO or effective diffusivity of aerosol particles), the validation exercise is highly relevant. They suggested the following correlation between turbulent diffusivity, area of room, and air change rate:

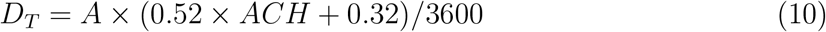

which is used for the present study as well. Recall that *A* is the area, *ACH* is the air changes per hour, therefore *a* = *ACH/*3600.

Denoting *τ* as the time duration of the event under considerations, the ensemble averaged number of infectious virions, generated from both speech and breath of the infected individual located at *x*_0_, *y*_0_, *z*_*o*_, that is inhaled at *x, y, z*_0_ upto time *τ* is denoted *𝒩*_*v*_(*x, y, z*_0_, *τ*) and it is given by:

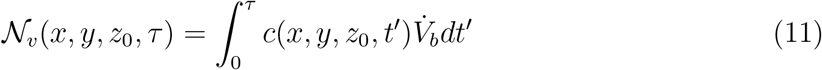

While infectious aerosol emissions from breathing take place over the entire time duration of the event, emissions from talking are assumed to occur only for a time *t*_*s*_ *< τ* (see supplementary material, section 3 for details). This is accounted for in our calculation of the corresponding time-varying virus concentration field. 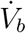 is the average volume of air inhaled per second. The local probability of infection due to infectious aerosols produced by speaking and breathing *𝒫*_*s*+*b*_(*x, y, z*_0_, *τ*) is calculated using the dose-response model originally proposed by Haas [42]. The dose-response constant is chosen as *r*_*v*_ = 1*/*1440 based on the estimations by Haas [43] and Schijven et al. [28] for the original SARS-CoV-2 variant. The subscript *s* + *b* indicates infection caused by exhaled aerosols due to speech (*s*) and breath (*b*) from the infected individual.

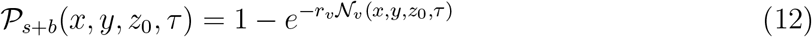

Th number of secondary infections *Z* within a room is thus given by

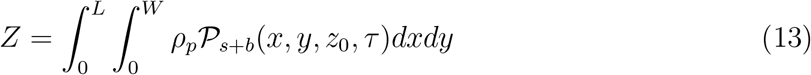

Here *ρ*_*p*_ is the susceptible population density (assumed to be uniform), estimated as *ρ*_*p*_ = *n/A* at the given point of interest (POI). POI is a term used in SafeGraph data-set and in Chang et al. [44] referring to a specific business location like a restaurant. *A* is the indoor area of that POI. In this paper, we place one infectious case at each POI. Therefore, *Z* is a measure of individual-level infectivity as well. Here *n* is the number of susceptible individuals present at a given POI i.e. *n* = *n*_*p*_ − 1 where *n*_*p*_ is the total number of people present at that POI. *N* is the sample space variable corresponding to *n*.

### 2.2 Algorithm for generating the pdf of secondary infections

With a basic semi-analytical framework available to compute the number of secondary infections inside a room of specified dimensions, for a given viral load, *ACH*, and occupancy, we use the algorithm detailed in Fig. 1 with randomized inputs from specified distributions (e.g., for viral load, air change rate) and available data (e.g., for occupancy, room area) to generate the pdf of 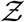.

The algorithm runs over *N*_*s*_ iterations where *N*_*s*_ = 103679 corresponding to the total number of full service restaurants in 10 US cities, available from SafeGraph data, used for generating a pdf at the end; *i* being the iteration index. First, the volume of the mucosalivary fluid ejected per unit time, evaluated from the exhaled respiratory droplet size volume size distribution (vsd) multiplied with a random sample from the log-normal viral load distribution, is fed into the code to generate the number of virions emitted per unit time. This is used in the turbulent diffusion solutions shown in Eqns. 4-7. Random samples of exposure time, speaking time, *ACH*, and area of POI, along with virus half-life, which is calculated separately, also act as input parameters for the solver. The resulting concentration field is used in the dose response model (with a specified dose response constant) to obtain probability fields. Next, probability fields are multiplied with the POI specific population density data (from SafeGraph) to obtain Z values. Once the index value reaches *N*_*s*_, the iteration ends and the pdf of 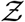 is calculated using *Z* values from every iteration. Note that the random variable - number of secondary infections at specific POIs or individual infectivity is denoted by *Z*, while the sample-space variable corresponding to *Z* is denoted by 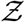. Pdf of 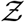 i.e. probability density function, defined as probability per unit distance in the sample space of 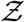 [35], is a powerful tool to describe overdispersion, irrespective of the specific number of events or samples. Furthermore pdfs are amenable to analytical descriptions. Fundamentally, any long tailed pdf represents an inherently overdispersed random variable, because the long tail represent finite (but could be small) probability of an event where *Z >>* mean of *Z*, while any pdf other than that represented by a delta function represents heterogeneity. In view of these, in this paper, the analysis of overdispersion will be addressed using both simulated and analytical pdfs of 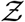.

Three key inputs - particle size distribution, viral load distribution, and occupancy information are described below, details of the rest of the inputs: ventilation rate distribution, speech time and exposure time distributions can be found in the supplementary material (section 3). A table summarizing the important parameter values, distributions, and sources can also be found there (Table 1).

#### 2.2.1 Particle size distribution of exhaled aerosols

The particle size distributions at the source of the expiratory events: speaking and breathing are obtained from the review by Pöhlker et al. [33]. A multimode lognormal fitting has been found to describe the corresponding distribution reasonably well. Number size distribution (nsd) for exhaled aerosol particles for different expiratory events can be described using a single function with event specific constants. The general form for the distribution can be expressed as

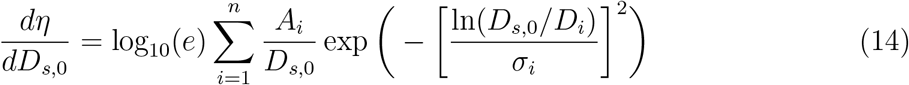

where *η* is the number concentration of particles (whereas *q*_*η*_ is the volume concentration of particles), *D*_0_ is the particle diameter at ejection i.e. at time *t* = 0, and *D*_*s*,0_ is the corresponding sample space variable. Moreover, *A*_*i*_ and *σ*_*i*_ are constants that depend on the mode and type of expiratory event. For further details the reader is referred to the supplementary material (section 1). Figure 2 shows the number and volume size distributions that are used in the simulation for the speaking and breathing events. The volume size distribution (vsd) is integrated over *D*_*s*,0_ to obtain the volume of the exhaled respiratory liquid per unit volume of air for the given expiratory event. This volume flow rate ratio multiplied with the individual viral load *ρ* and the volume flow of air per unit time, yields the ejected number of virions (RNA copies) emitted per unit time i.e. the source term 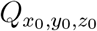 of Eqn. 4. Here we choose particles only with *D*_*s*,0_ *<* 100*µm* as the larger ones will settle in less than 10 s even after accounting for evaporation [19]. A detailed description of the particle deposition velocity calculation is presented in the supplementary material (section 2).

**Figure 2:**
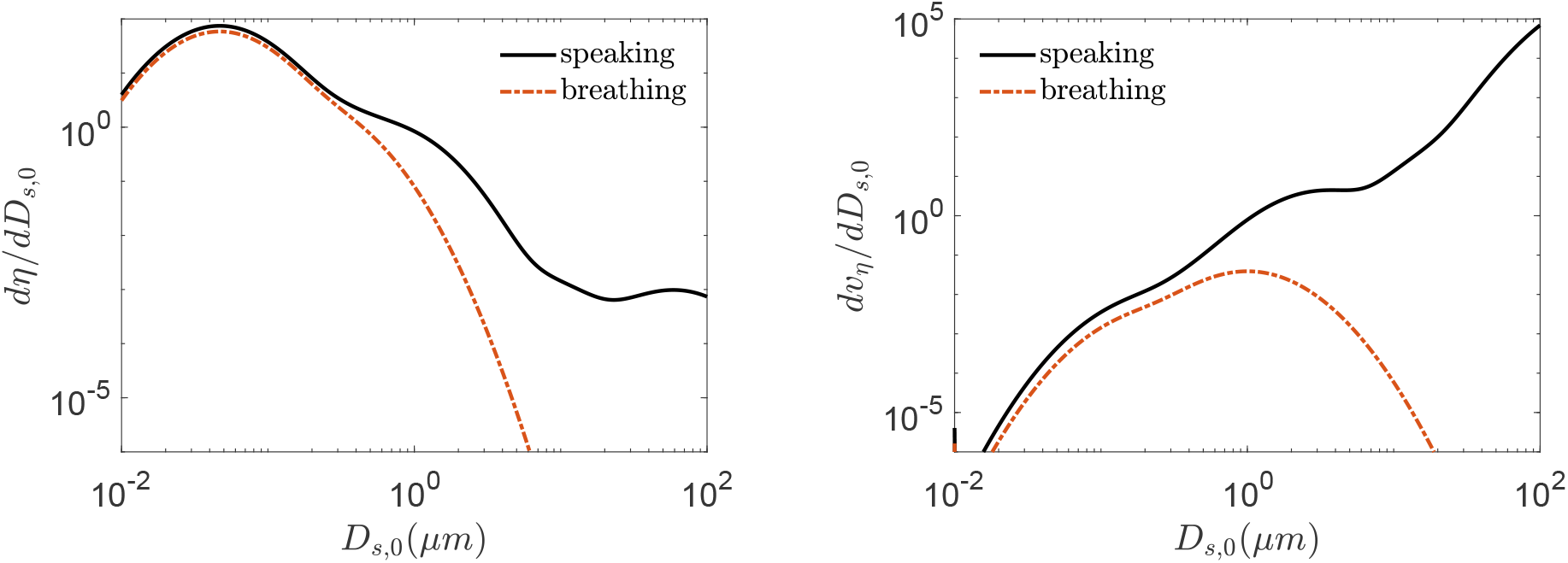
Aerosol number size distribution (nsd) on left, and volume size distribution (vsd) on right, for speaking and breathing as expressed by Eq. 14 and Eq. 4 respectively, from [33] as a function of initial particle diameter sample space variable *D*_*s*,0_ i.e. at the moment of exhalation.

#### 2.2.2 Viral load distribution

Viral load, (*ρ*) has been associated with infectivity and thus, the number of secondary infections for a given setting [45, 46]. Analyzing respiratory droplet and aerosol laden cough jets Chaudhuri et al. [19] showed that the corresponding number of infected individuals could vary by orders of magnitude due to variation in the viral load. Chen et al. [10] analyzed a large number of SARS-CoV-2 viral load databases and suggested that viral load is an important contributor to heterogeneity in secondary infections. They also showed that the viral load distributions were similar for symptomatic and asymptomatic stages of infection. This point was further amplified by direct measurements of Yang et al. [47] who showed that viral load distributions are nearly identical among hospitalized (symptomatic) and asymptomatic population. In this paper, we utilize measurements of viral load in asymptomatic (including presymptomatic) population from Yang et al. [47] as an input into the algorithm shown in Fig. 1. According to [47] at the time of saliva collection, the infected individual was either asymptomatic or presymptomatic. The pdf of *ln*(*ρ*_*v*_) (*ρ*_*v*_ is the sample space variable of *ρ*) is shown in Fig. 3. We generate and use samples of *ρ* from this log-normal distribution in our calculations.

**Figure 3:**
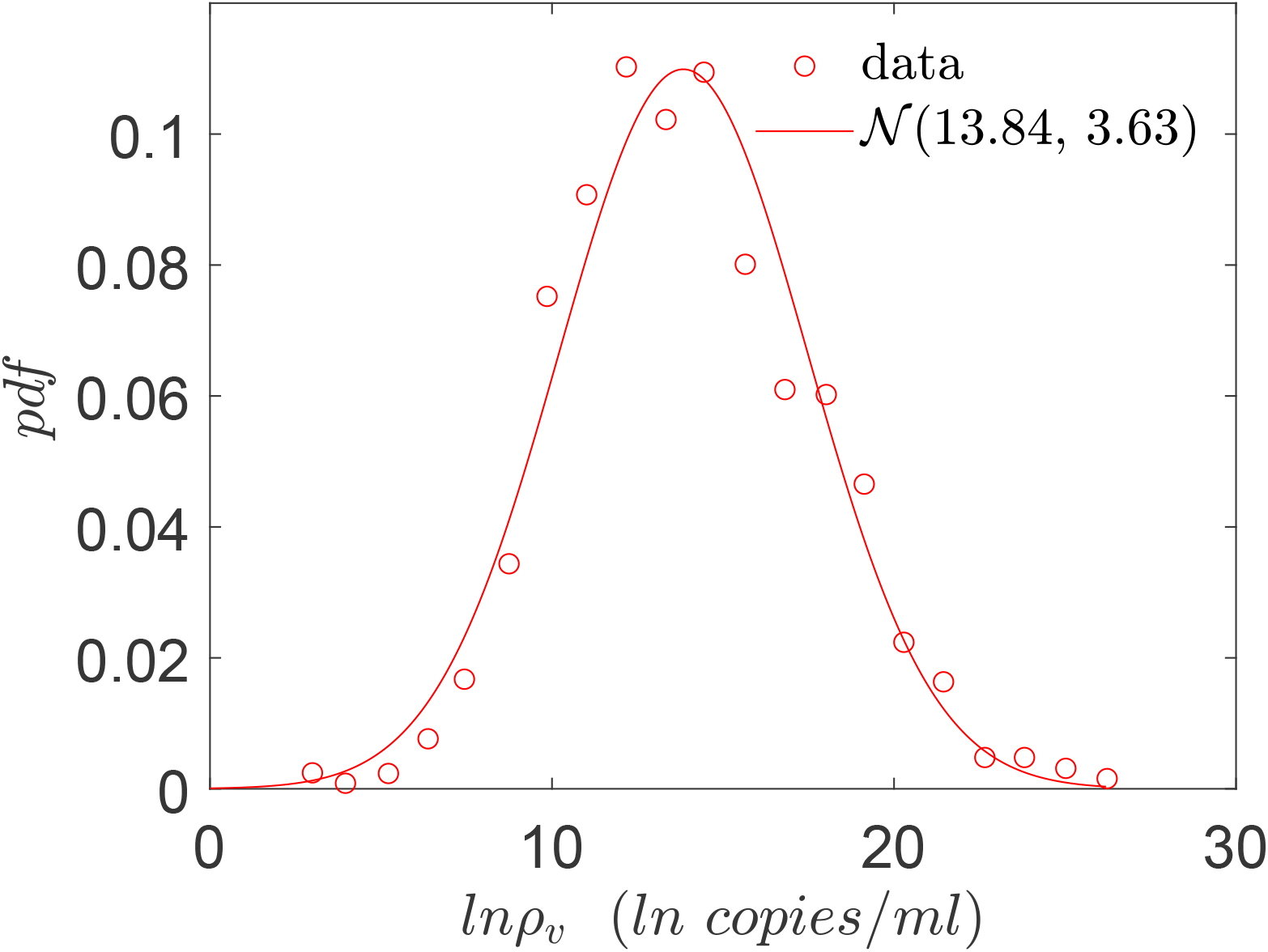
Viral load pdf for asymptomatic (including presymptomatic) population, based on the histogram data from Yang et al. [47]. The red line shows a normal distribution with *µ* and *σ* given by 13.84 and 3.63 respectively.

#### 2.2.3 Occupancy and area of different points of interest from SafeGraph data

Two of the important inputs in the simulation are the areas of different points of interests (POI) and the number of people occupying them during each time period of interest. These data were obtained from SafeGraph - a company that collects anonymous data from mobile devices. For our simulation, data that is available from individual confined spaces, rather than that from a collection of several indoor spaces, was felt to be most appropriate. Hence, we used SafeGraph data for areas of full service restaurants (POI) over ten cities in USA, namely Atlanta, Chicago, Dallas, Houston, Los Angeles, Miami, New York City, Philadelphia, San Francisco, and Washington D.C. The SafeGraph-tabulated area for each restaurant is multiplied by 0.5 to convert the total area of a given restaurant to the corresponding sitting area since the dining area is estimated as 50 % of the total restaurant area (based on typical restaurant design guides [48]; change of this factor does not qualitatively change the observations). The occupancy information in these POIs between hours 12:00 - 13:00 and 18:00 - 19:00 over seven days starting from March 1, 2020 is obtained from the data sets created by Chang et al. [44] who developed a mobility network based SEIR model using the SafeGraph data. These two time periods represent typical lunch and dinner times, and hence highest occupancy periods of any day. The pdf of the averaged number of susceptible individuals in restaurants (total occupancy minus one) between hours 12:00 - 13:00 and 18:00 - 19:00 over ten US cities is shown in Fig. 4. Also, shown in the figure is an exponential fit and the corresponding fitting parameter.

**Figure 4:**
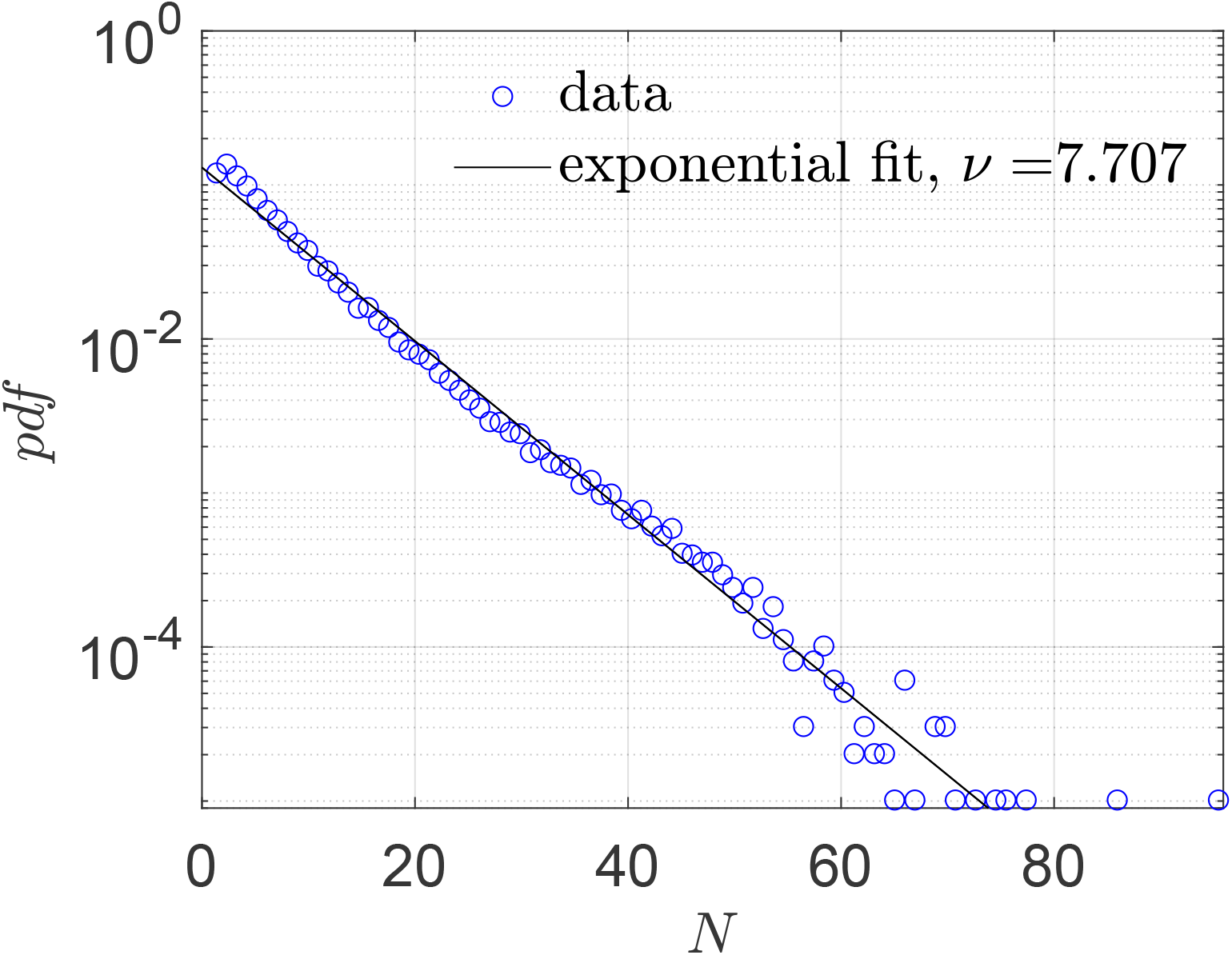
Pdf of number of susceptible individuals in full service restaurants at ten US cities, from SafeGraph data.

## 3 Results and discussion

### 3.1 Spatial distributions of particle concentration and infection probability for fixed conditions

First, we present the spatial distribution of the virus concentration (RNA copies/*m*^3^ of air) with the source at *x* = 2.5*m* and *y* = 2.5*m* from the origin (at the bottom left corner) of a 10m x 10m x 3m room after 15 minutes of aerosol exhalation by speech and breath. In Fig. 5 the first row (a-c) presents results with a constant viral load *ρ* = 10^9^ copies/ml, but with increasing *ACH*. Note that the third column represents a case without wall reflections and hence can simulate outdoor conditions. The second row of Fig. 5 (d-f) represents a constant but five times higher viral load of *ρ* = 5 × 10^9^ copies/ml. The results demonstrate strong inhomogeneity of virus concentration and also show that contours scale linearly with *ρ* for the same *ACH* due the linear nature of the governing Eqn. 3. Using the dose response model (Eqn. 12) the corresponding contours of probability of infection are shown in Fig. 6. Indeed near unity *𝒫*_*s*+*b*_ are found near the source, and there is decay with distance from the source. The *𝒫*_*s*+*b*_ contours do not scale linearly with *ρ* due to the nonlinear nature of the dose response model. Increasing *ACH* invariably reduces virus concentration for a given *ρ*. However, the reduction in the probability of infection may not be proportional to the reduction of the virus concentration due to the non-linearity involved in the dose response. Interestingly, the simulated outdoor conditions (c, f) show much smaller infection probability both near and far from the source w.r.t. the confined cases. This can be attributed to the inherently higher *ACH* inside the volume of interest, but primarily due to absence of confinement which allows the virus concentration to freely decay with space.

**Figure 5:**
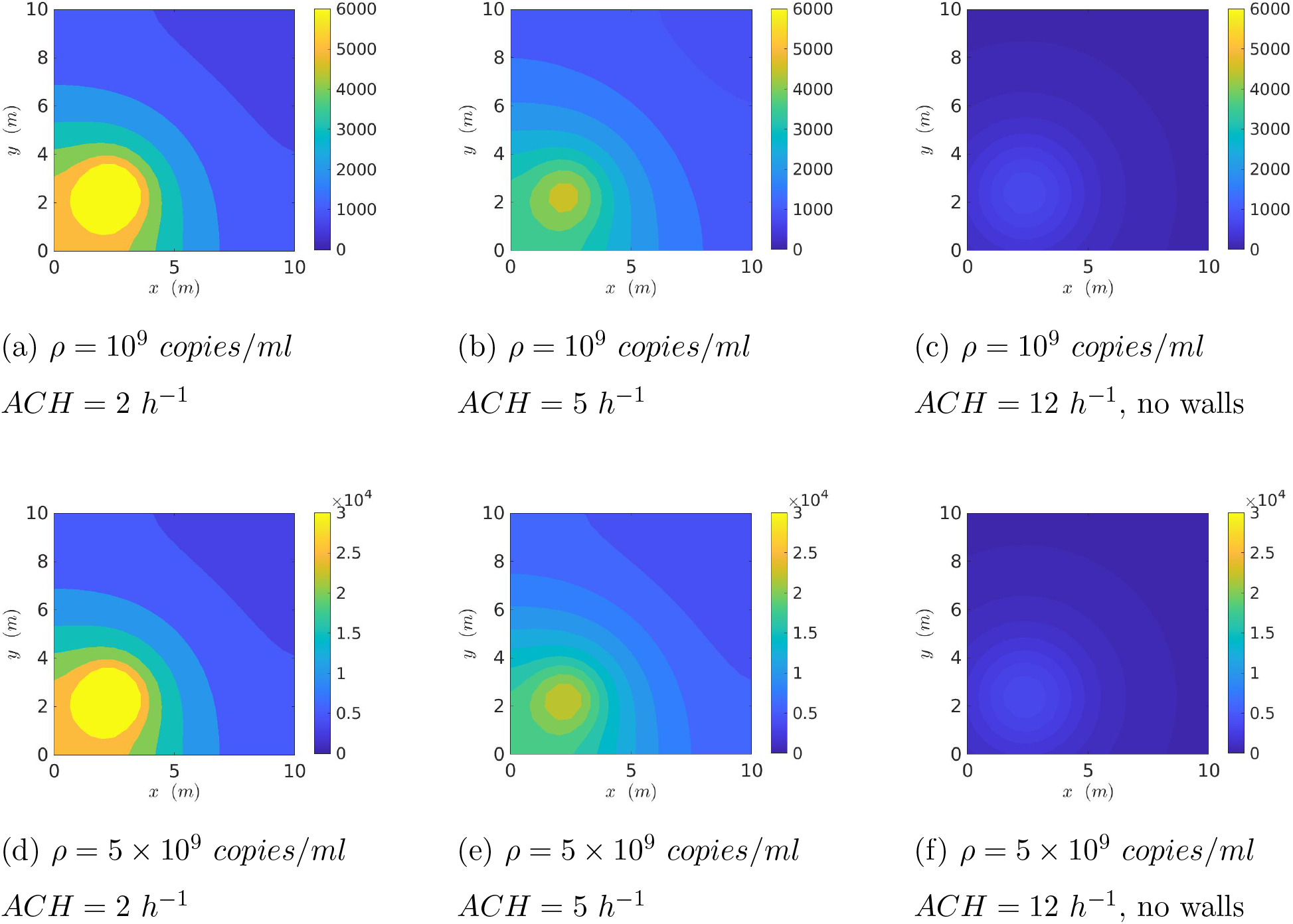
Contour plots of spatially resolved virus concentration (RNA copies/*m*^3^) at time *t* = 15 minutes from start of expiration event (source located at *x* = 2.5*m, y* = 2.5*m*).

**Figure 6:**
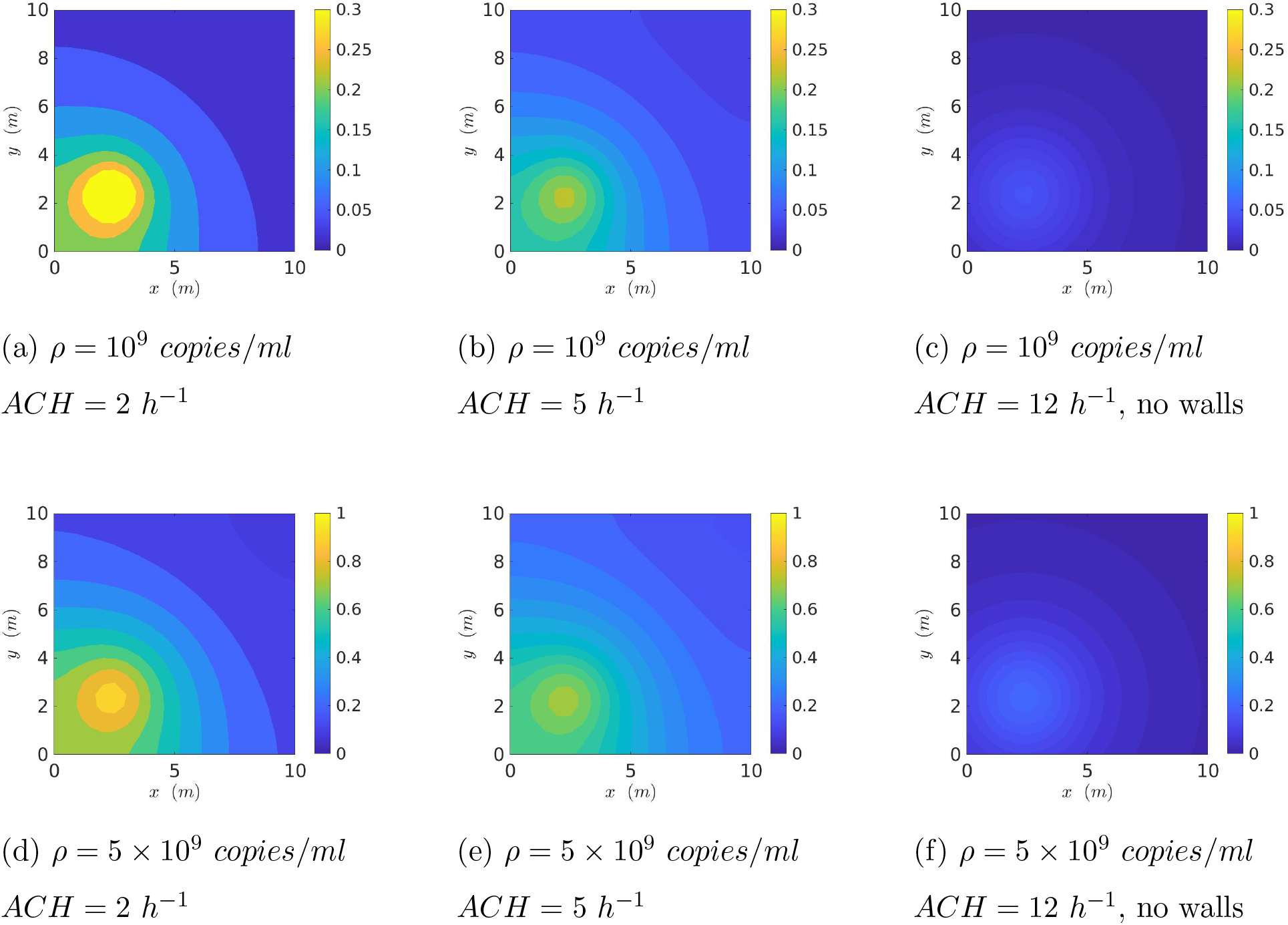
Contour plots of spatially resolved probability of infection *𝒫*_*s*+*b*_(*x, y*) at time *t* = 15 minutes from start of expiration event (source located at *x* = 2.5*m, y* = 2.5*m*).

### 3.2 Statistical distributions of secondary infections generated from realistic input distributions

A simulation based on the algorithm presented in Fig. 1 is run for each data point available from the predefined SafeGraph dataset, resulting in a sample size of *N*_*s*_ = 103679. We place one infected individual at each POI, at random locations within its premises. As such, most inputs, including viral load, *ACH*, exposure time, speaking time, source location (*x*_0_, *y*_0_), are randomized. *ACH* is generated from a log-normal distribution with a median *ACH*= 2.16 hr^−1^, such that *µ*_*ACH*_ = 0.7701, *σ*_*ACH*_ = 0.7554 based on measurements by Bohanon et al. [49]. Room height *H* = 3 m, height of source (seated) *z*_0_ = 1 m, indoor conditions (*T* = 21.7^*o*^*C, RH* = 0.50, *UV*_*index*_ = 0), and dose response constant *r*_*v*_ = 1*/*1440 are held constant. The resulting distribution of number of secondary infections - pdf of 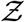 is presented in Fig. 7. An analytical solution 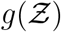 to be derived in sub-section 3.3 is also shown in the figure.

**Figure 7:**
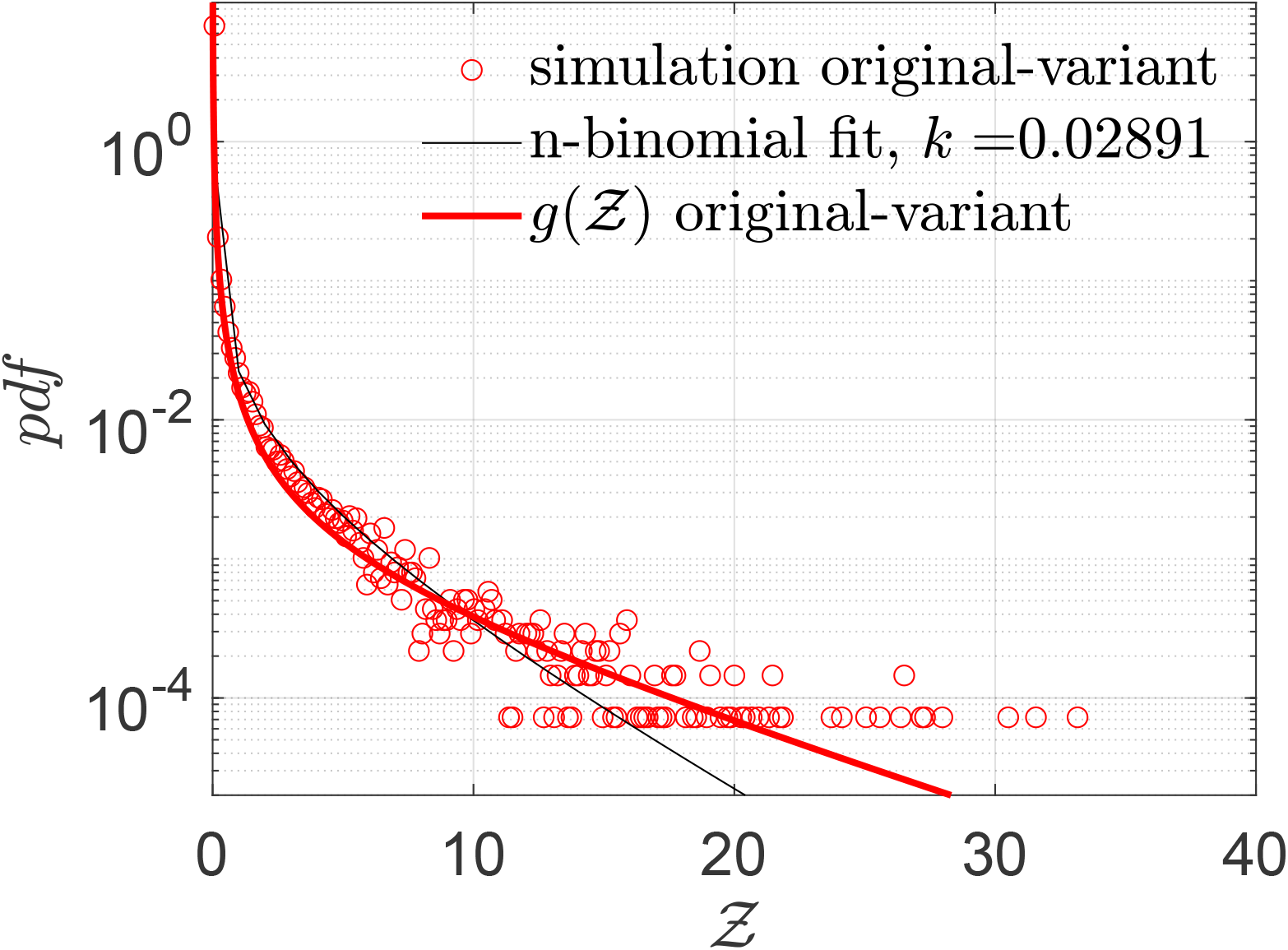
Pdf of the number of secondary infections: 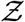 and negative binomial fit. The analytical function *g* is given by Eqn. 22. *g*(*µ* = 13.84, *σ* = 3.63, *α* = 1.123×10^−10^, *ν* = 7.71).

The stretched tailed nature of the simulated pdf is immediately apparent. This shows that there is small but finite probability of tens of secondary infections per infected individual. For this simulation the *mean*(*Z*)_*s*_ = ⟨*Z*⟩_*s*_ = 0.14 indicating that over this one hour, on average less than 1 person got infected per infector. The calculated total number of infections is 14482, with many of the infections occurring in large superspreading events. As such, only 3.57 % of the infected individuals infected 80 % of the population over this time. This could also be the reason why it is generally difficult to culture the virus from the air, though that was unequivocally demonstrated by Lednicky et al. [50]. High probability of infection, which as shown in the paper typically occurs at high viral load, could be correlated with high probability of virus detection in the air. Direct virus detection from air could therefore necessitate sampling from a large population of infected individuals. Clearly, the finite probability of *Z >>* ⟨*Z*⟩ recovers the inherently overdispersed nature of SARS-CoV-2 transmission dynamics. Fitting a negative binomial probability distribution function to the 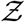-pdf yields a good fit, with a dispersion parameter *k* = 0.029. While the fit quality worsens at the pdf tails, the dispersion parameter is in the same order as the corresponding values for SARS and measles estimated by Lloyd-Smith et al. [1]. However, it is to be noted that we are considering only infections over a period of about 1 hour on average, as opposed to the entire course of infection, hence ⟨*Z*⟩ should not be interpreted as *ℛ*_0_. Similarly, the qualitative *k* value thus obtained should be interpreted with care.

Figure 8 shows the joint pdf of *ρ* and 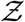. The close correlation of the two random variables across nearly six orders of magnitude is immediately apparent from this figure. While correlation may not imply causality in general, it is reasonable in this case that the extreme variation in *ρ* is causing a similar variation in *Z*, with other parameters controlling the slope and strength of the correlation.

**Figure 8:**
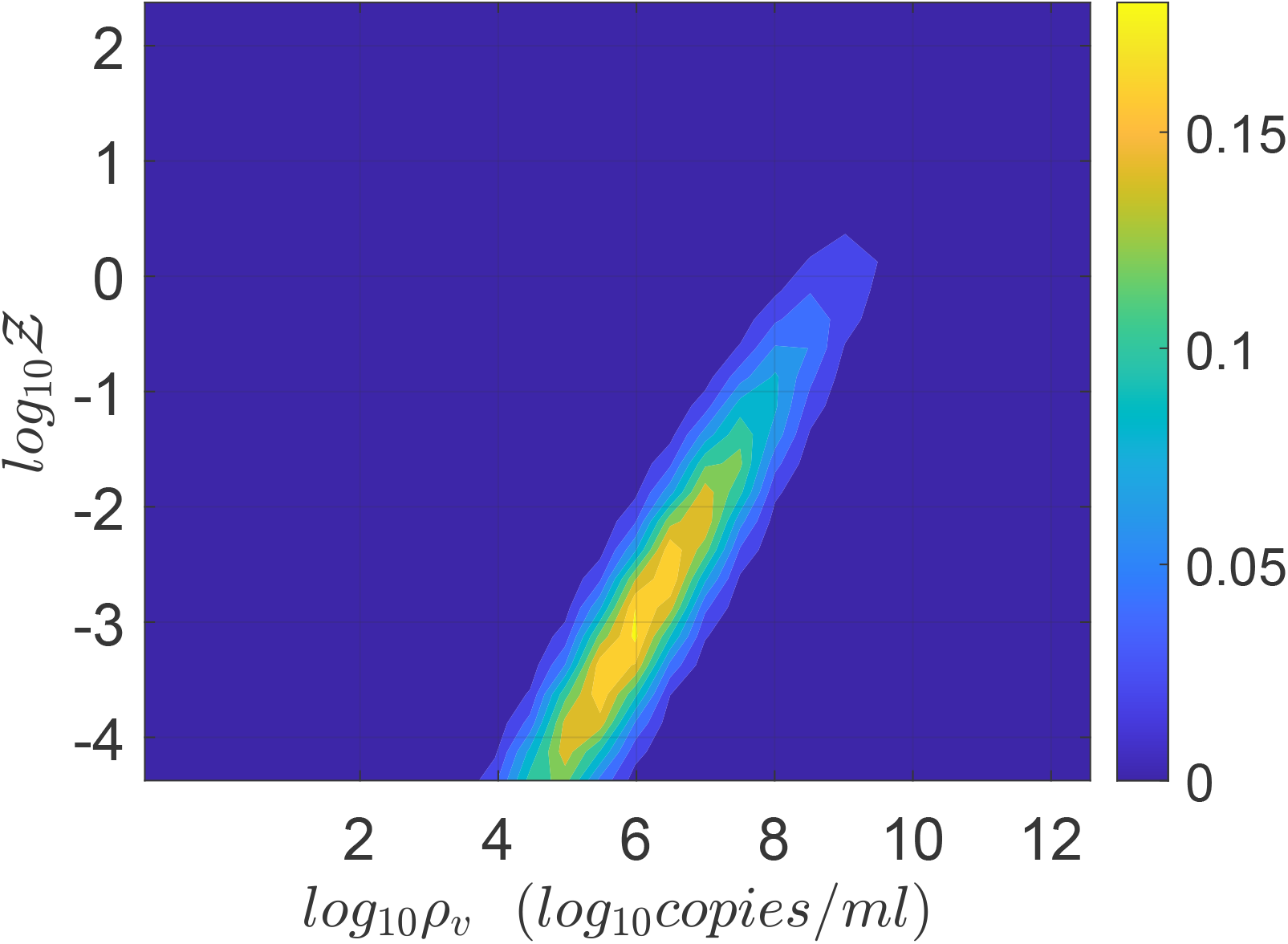
Joint pdf of viral load and number of secondary infections.

We present the jpdf of *ACH* and *Z* in Fig. 9. It is apparent that the highest number of infections *Z* occur at lower air exchange rates, as expected; however, the majority occur at an intermediate air exchange rate of about 1-1.5 *ACH*, in part because very low and very high air exchange rates are simply less common. Furthermore, it is also clear that dispersion of *Z* and *ACH* are indeed negatively correlated. The effect of universal, high ventilation rates, and masks will be taken up later.

**Figure 9:**
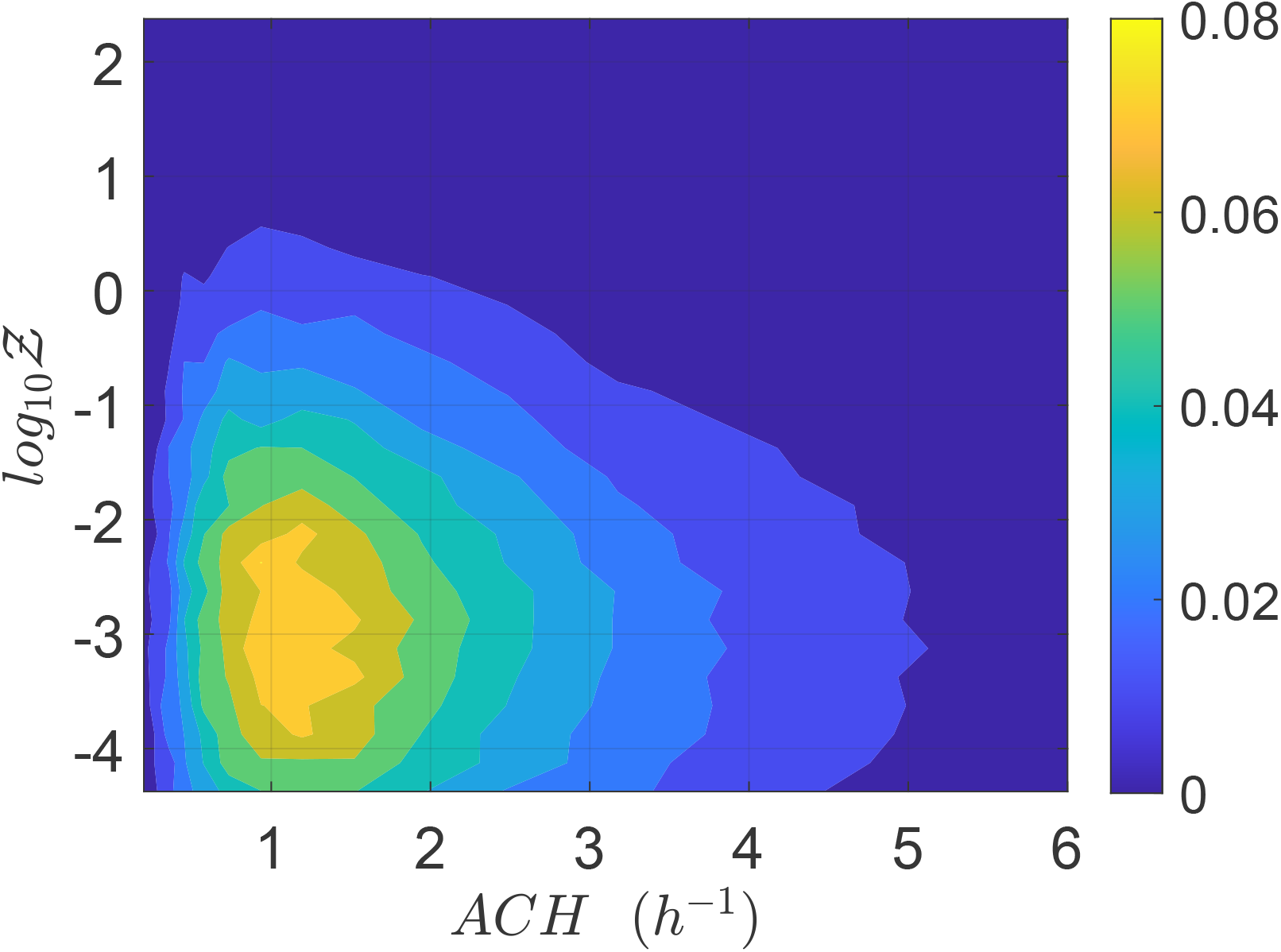
Joint pdf of *ACH*and *Z*.

### 3.3 Derivation of the analytical pdf of 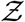: 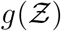

The viral load pdf from Yang et al. [47] can be well approximated by a log-normal distribution with parameters *µ* and *σ* given by *µ* = 13.84 and *σ* = 3.63. This is shown in Yang et al. and also shown in Fig. 3. As such, we used the following pdf of viral load *f* (*ρ*_*v*_) for generating inputs into the simulation

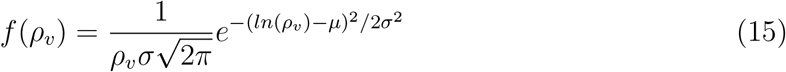

Due to its extremely large (over *𝒪*(12)) variation, our analysis shows that viral load *ρ* is one of the most dominant variables in controlling overdispersion of secondary infections *Z*, as is apparent from Fig. 8. This result can also be presented in terms of secondary attack rate - a more generalized descriptor, defined as 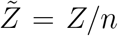 where *n* is the number of susceptible individuals present at the given POI over the period of interest *τ*. 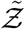 is the sample space variable corresponding to 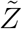. Variation of 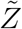 w.r.t. *ρ* is shown in Fig. 10. Clearly, this plot reflects the dose-response function (Eqn. 12) on a macro scale given the dominant influence of *ρ* in controlling probability of infection and eventually secondary attack rates. Therefore, we propose a function similar to Eqn. 12 to model the response of 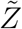 to *ρ* variation. This is shown below:

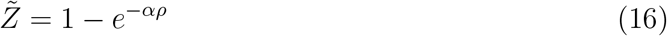

**Figure 10:**
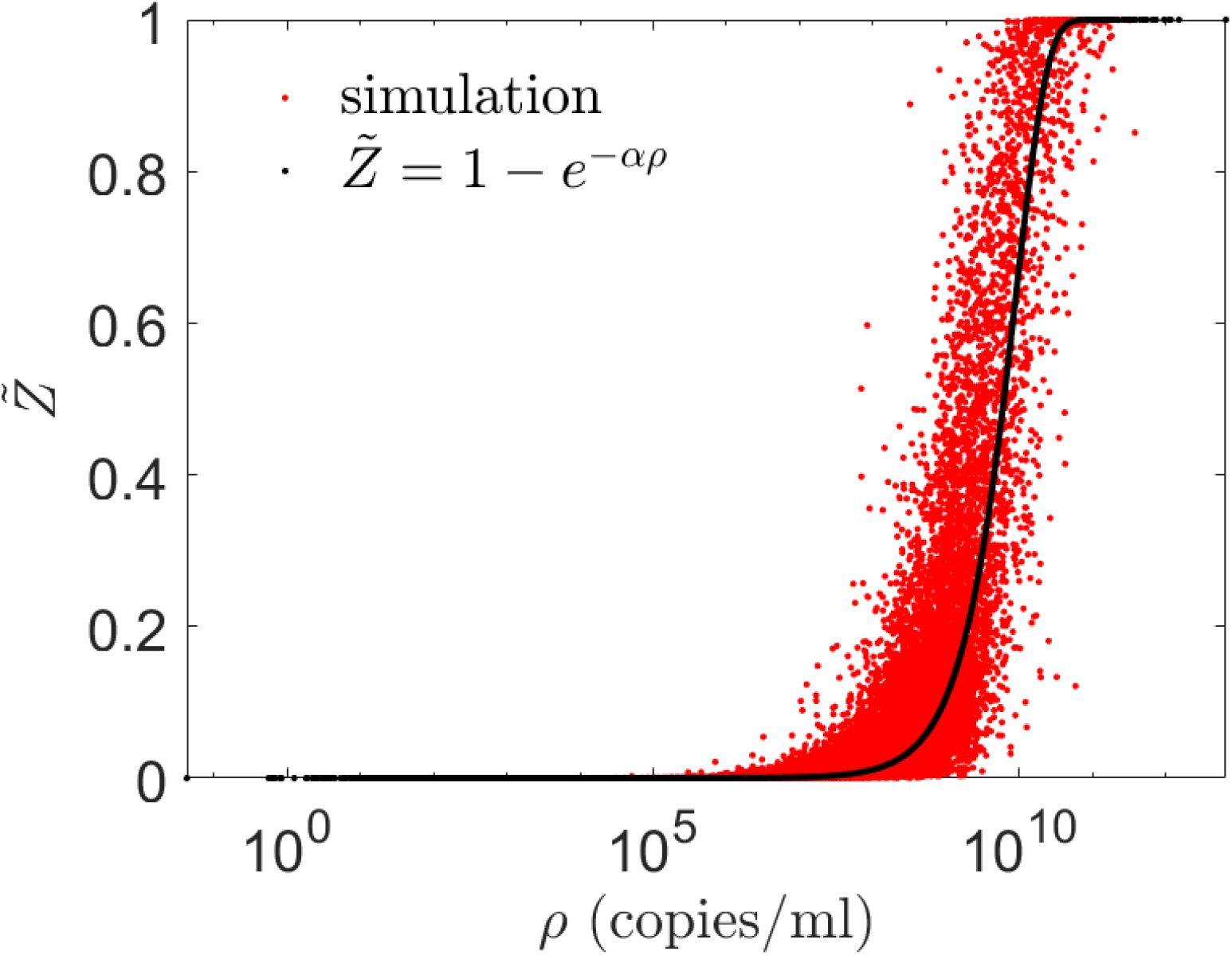
Scatter plot of viral load (*ρ*) vs secondary attack rate 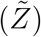. The black dot curve shows Eqn. 16 with *α* = 1.123 × 10^−10^ ml/copies.

We can also write

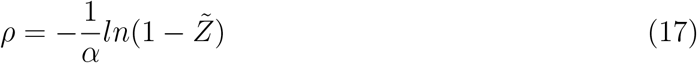

The constant *α* can be estimated as the inverse of the average number of virions inhaled per unit volume of mucosalivary liquid ejected that is required for infection probability of 1 − *e*^−1^ = 0.63. For the average room volume ⟨*V* ⟩ = 6.34 × 10^2^ m^3^, average speaking time ⟨*t*_*s*_⟩ = 1469 s, average exposure time ⟨*τ* ⟩ = 3914 s, average air change rate, ⟨*ACH*⟩= 2.87 hr^−1^, and deposition parameter based on average room area and volume *β*_0_ = 0.002 s^−1^, *α* can be estimated as

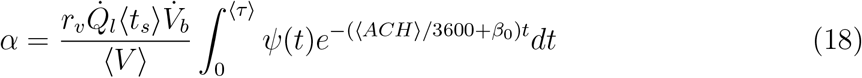

*ψ*(*t*) is given by Eqn. 9, and 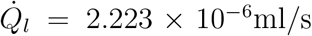. Note that all the ⟨ ⟩ quantities mention averages over the distributions used in the present simulation. Equation 18 yields *α* = 1.123 ×10^−10^ ml/copies. The comparison of Eq. 16 and the simulation results are shown in Fig. 10.

With the functional form of the pdf of the viral load known (given by Eqn. 15) we can immediately substitute Eqn. 17 into Eqn. 15 to eventually derive the pdf of 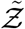 using the generalized equation below.

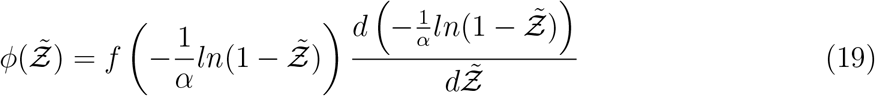

Using the log-normal form of *f* we derive the analytical function below which could be used to model the pdf of secondary attack rate 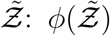. However, the same method should be applicable to other functional forms of *f*, like a Weibull distribution as in [10] instead of log-normal.

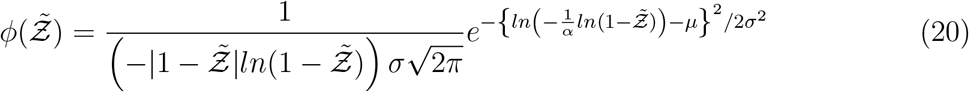

Comparison of Eqn. 20 with the simulation results is shown in Fig. 11.

**Figure 11:**
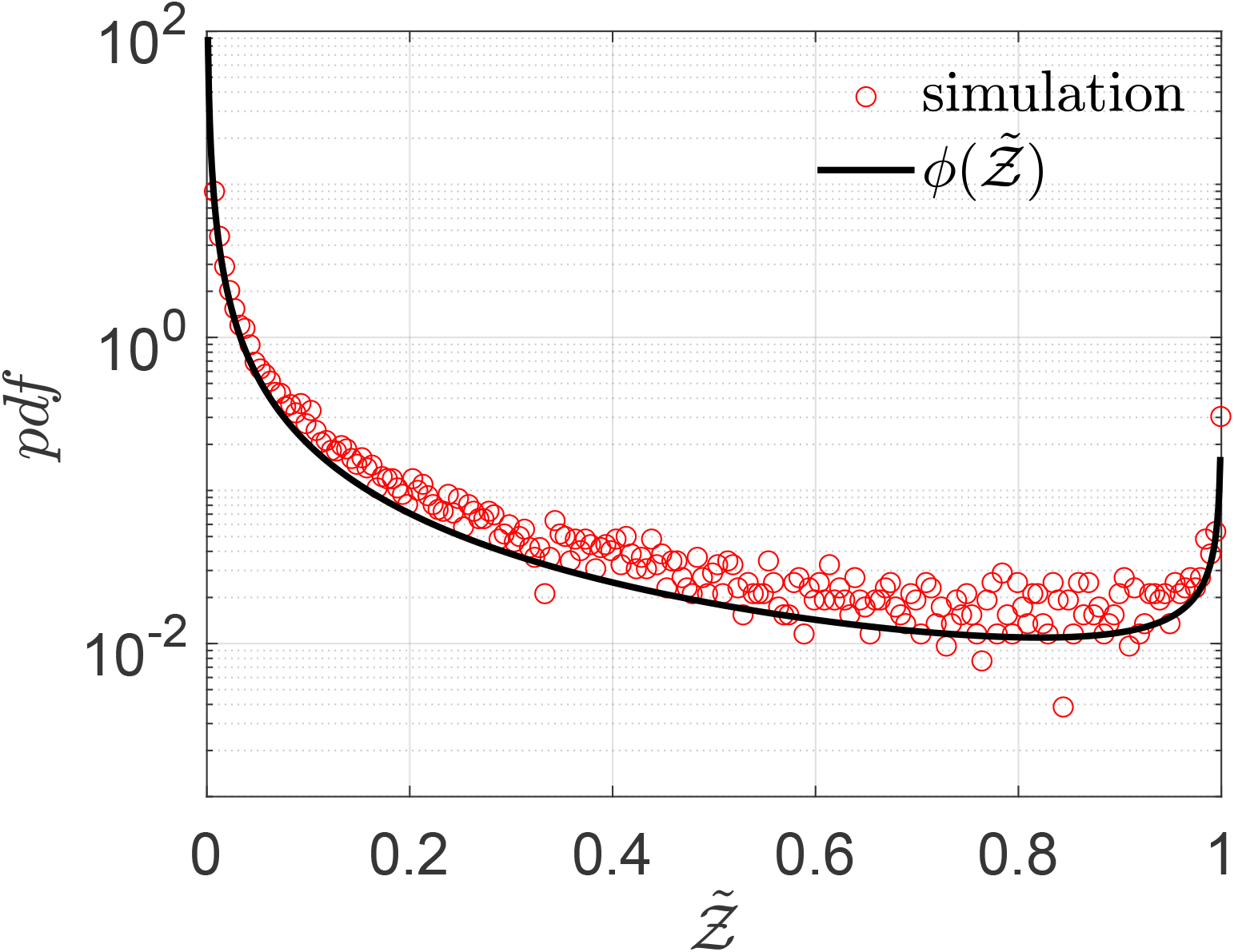
Pdf of 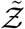 and its comparison with the analytical result given by Eqn. 20

It is evident that Eqn. 20 describes the simulation data to good quantitative accuracy. It is also remarkable that very important effects of area, *ACH*, virus kinetics, exposure and speaking times could be encapsulated within one constant *α*. It is to be recognized that the equation is valid only for 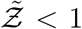. This is an inherent feature emanating from the derivative of the functional form of the dose response model which yields the 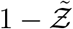 term in the denominator. Importantly, the equation can describe the range 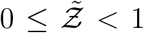 with high degree of veracity. Now, we can write *Z* = *N* (1 − *e*^−*αρ*^) using Eqn. 16. For a fixed *α*, clearly *N* and 1 − *e*^−*αρ*^ are independent random variables. Therefore, for any given pdf of *N* given by *h*(*N*), pdf of 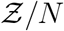 given by 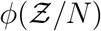 and a known *α*, and using the general equation that describe the pdf of the product of two independent random variables, we can write the pdf of 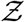 given by

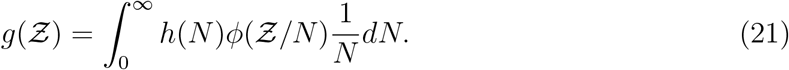

Using the exponential distribution 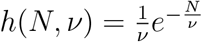 shown in Fig. 4 we derive for 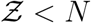

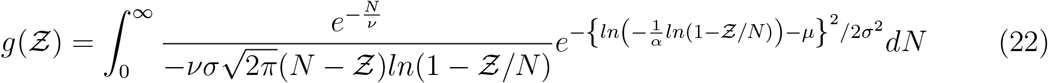

It is to be noted that the constants *µ, σ* are properties of the viral load distribution, *ν* is the constant of the occupancy distribution, and *α* encapsulates 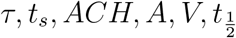 etc. according to Eqn. 18. It is apparent that the pdf 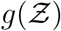 is stretched to higher (lower) 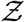 values when *µ, σ, ν* or *α* increases (decreases). The remarkable match between this analytical function: *g* - the analytical pdf of the number of secondary infections and that obtained from the simulation data has been shown in Fig. 7. We revisit it here for further discussion. It is evident that the stretched tail of the pdf of 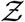 results from the two stretched exponential functionals of 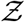 and *N*. This can be verified by noting that when either *σ →* 0 or *ν →* 0 the overdispersion of *Z* vanishes. The first stretched exponential arising from the lognormal distribution of viral load *ρ*_*v*_ and the latter from the exponential distribution of number of people at the different POIs. Therefore, it is the joint contribution of overdispersed viral load and overdispersed occupancy that results in overdispersion of secondary infection numbers causing superspreading events. This is shown here with a single equation. The ⟨*Z*⟩ obtained from the analytical pdf, given by 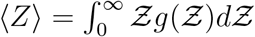. We find mean and standard deviation as ⟨*Z*⟩= 0.13, *std*(*Z*) = 0.99, respectively, in comparison to ⟨*Z*⟩_*s*_ = 0.14 and *std*(*Z*)_*s*_ = 0.87 from the simulations. The analytical pdf 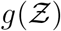 is expected to be a generalized result and could be applied for any large number of indoor POIs, without much restriction on their type.

### 3.4 Variants and mitigation measures

Finally, we test whether the derived Eqn. 22 can describe overdispersed transmission associated with a different viral load distribution equally well. To this end, we generate a viral load distribution with a mean which is 1000 times the mean of the original distribution presented in Fig. 3. Thus, while Fig. 3 represents the pdf of the original variant, the new log-normal distribution characterized *µ* = 20.67 and *σ* = 3.63 might represent the pdf of viral load of the *δ*−variant infected individuals. Early data from Li et al. [51] suggest that the viral loads of the *δ*−variant could be as high as “1000 times greater compared to A/B lineage infections during initial epidemic wave in China in early 2020”. The simulation results in terms of 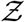-pdf are presented in Fig. 12. The greater transmissibility of the *δ*−variant under the assumption of 1000-fold higher mean viral load is immediately apparent. In comparison to the ⟨*Z*⟩= 0.13 for the original variant, the ⟨*Z*⟩= 2.64 for *δ*. Therefore, just based on viral load, according to the calculations and model, *δ*−variant could be nearly 20 times higher transmissible on average w.r.t. the original variant, over about an hour of contact. Interestingly, with just increased *µ*, Eqn. 22 can capture the pdf of 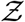 for the *δ*−variant, remarkably well, alongside the one for the original variant. However, it is to be recognized that there is some uncertainty on the assumption that mean viral load of the *δ*-variant is 1000 times higher than the original variant. Data also suggest that viral load and infectiousness potential (using a proxy of culture-positivity for example), while monotonic in nature, may not demonstrate a linear relationship [52]. Furthermore, ⟨*Z*⟩ ≠ *ℛ*_0_. Hence, the enhancement factor thus found is valid only within the context of the assumptions and scope of this work.

**Figure 12:**
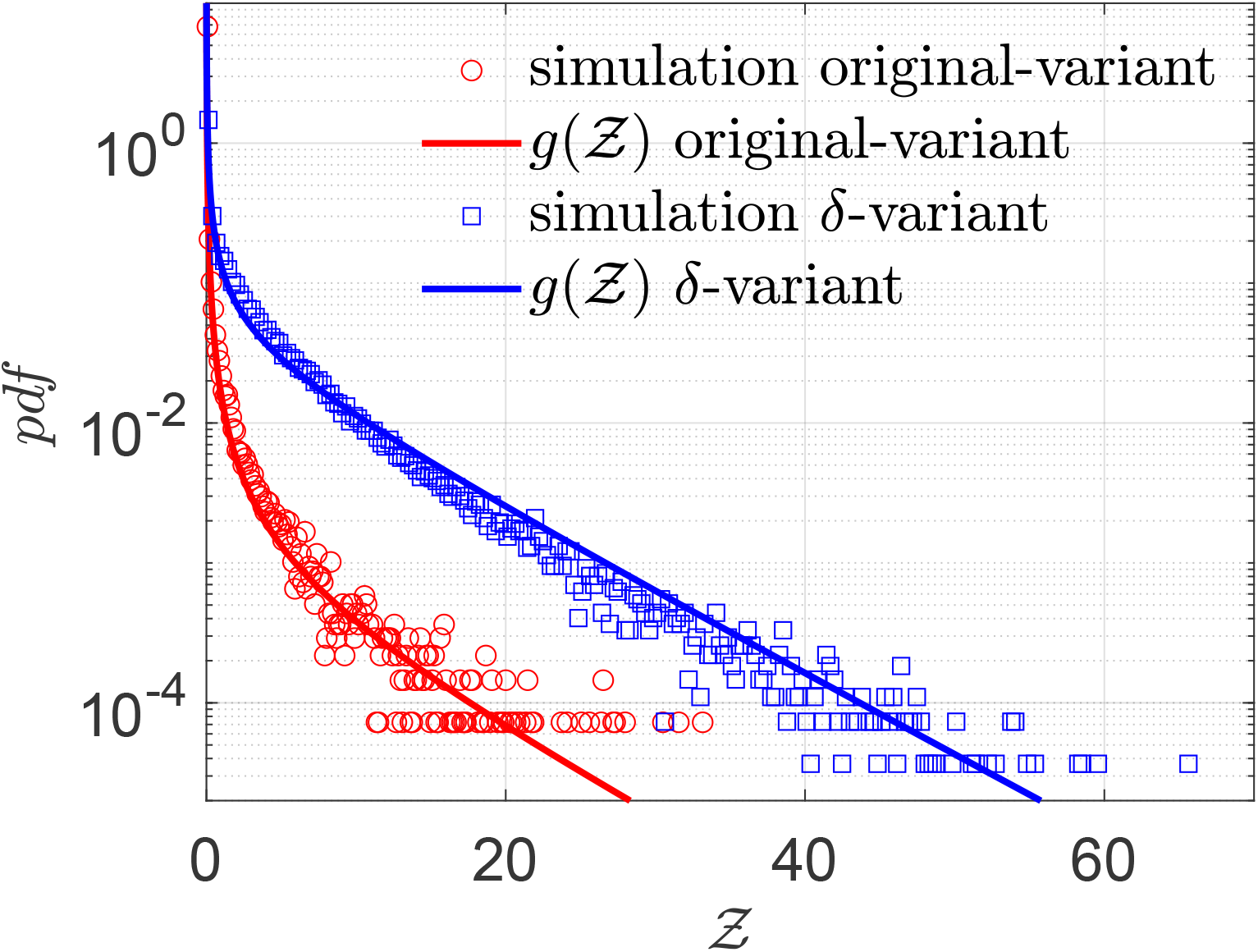
Pdf of 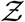 and its comparison with the analytical result given by Eqn. 22 for original variant (red circle symbols, red bold line) *g*(*µ* = 13.84, *σ* = 3.63, *α* = 1.123 × 10^−10^, *ν* = 7.71) and for *δ*−variant (blue square symbols, blue bold line) *g*(*µ* = 20.67, *σ* = 3.63, *α* = 1.123 × 10^−10^, *ν* = 7.71).

Finally, we explore the effect of adopting uniformly high ventilation rates and masks, on the distribution of secondary infections. To that end, we keep the ventilation rates constant at *ACH* = 5 hr^−1^ and assume that the entire population (including the infectors and susceptibles) are wearing masks that provide 50% reduction by volume of exhaled aerosols and 50% reduction in the correspondingly inhaled aerosols. The results are shown in Fig 13.

**Figure 13:**
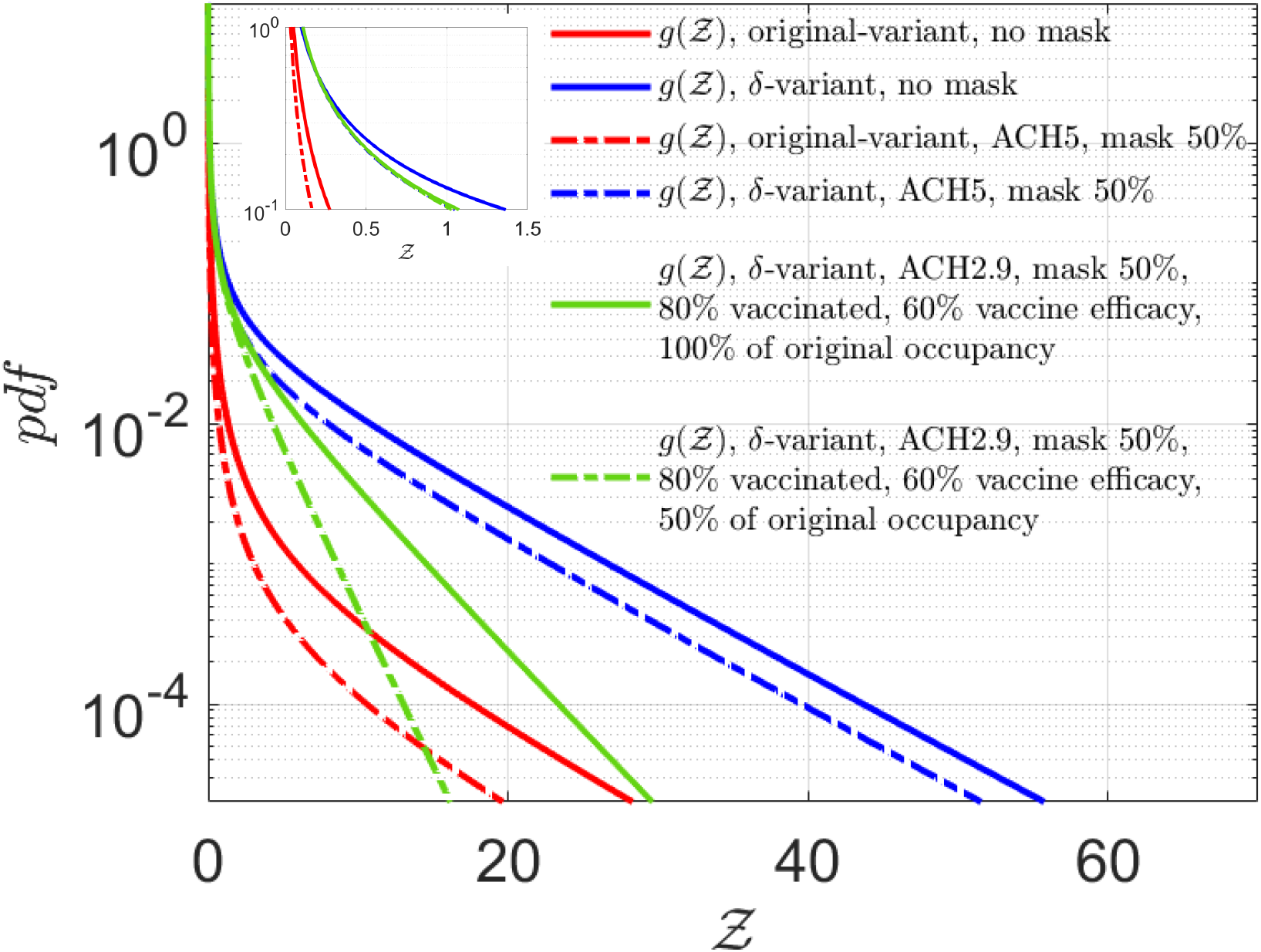
Effect of masking, fixed ventilation rates, vaccines, and reduced occupancy. Solid red and blue lines correspond to the analytical solutions given by Eqn. 22 for the original and *δ*−variant respectively, for ⟨*ACH*⟩= 2.9 hr^−1^ and without masks, as shown in Fig. 12. For fixed *ACH* = 5 hr^−1^, and with masks blocking 50 % volume of aerosols during inhalation and exhalation, the dashed red line shows the analytical solution for original variant *g*(*µ* = 13.84, *σ* = 3.63, *α* = 2.36 × 10^−11^, *ν* = 7.71) while the dashed blue line shows the analytical result for *δ*−variant *g*(*µ* = 20.67, *σ* = 3.63, *α* = 2.36 × 10^−11^, *ν* = 7.71). The solid green line: *g*(*µ* = 20.67, *σ* = 3.63, *α* = 2.81 × 10^−11^, *ν*_*v*_ = 4.01) shows the effect of 80% vaccination coverage, with 60% vaccine efficacy, with all individuals wearing masks that block 50% of the aerosols during exhalation and inhalation. The dash-dotted green line: *g*(*µ* = 20.67, *σ* = 3.63, *α* = 2.81 × 10^−11^, *ν*_*v*_ = 2.00) shows the effect of 80% vaccination coverage, with 60% vaccine efficacy, with all individuals wearing masks that block 50% of the aerosols during exhalation and inhalation along with occupancy restriction to 50% of the original occupancy. The top left inset shows the zoomed in view of the left side of the pdfs. Please refer Table 3 in the supplementary material for detailed values.

We observe that such interventions result in significant reduction in transmissibility for the original variant; reducing the mean to ⟨*Z*⟩= 0.04 (from ⟨*Z*⟩= 0.13 obtained without any interventions) with significant reduction in the extension of the tail. Such intervention effect remains substantial, yet less pronounced for the *δ*-variant where the ⟨*Z*⟩= 1.69 (w.r.t. ⟨*Z*⟩= 2.64 obtained without any interventions) and tail remains sufficiently stretched with some shift in the overall pdf towards lower 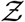. Once again, we note that these numbers are obtained over nearly an hour of exposure time on average. Note that, in our model, *α* does not change between the original and the *δ* variant. Only the *µ* increases, resulting in an increase in the higher proportion of secondary attack rates close to unity as the virus strain switches from the original variant to the *δ*−variant. A more detailed analysis of the influence of individual parameters: mean viral load, mean occupancy, and mean ventilation rates on the mean and standard deviation of *Z* could be found in the supplementary material (section 5).

Within the scope of the present study - social gatherings in restaurants, we ask what kind of spread could be expected for the *δ*−variant given the period of exposure and available occupancy data, in a population where a large fraction is already vaccinated? This is shown in Fig. 13 by the green curves. Using the realistic *ACH* distribution and with masks that can reduce both emission and inhalation of aerosol volumes by 50%, respectively, we do a basic calculation including the effect of vaccination. Assuming vaccine efficacy *η*_*vac*_ = 0.6 and vaccination coverage efficiency *η*_*cov*_ = 0.8 representing fraction of the population vaccinated, we estimate the new population of susceptible individuals at a given POI as *n*_*v*_ = (1 − *η*_*vac*_*η*_*cov*_)*n*. We do not consider any change in viral load or change in distribution of infectious cases. Fitting an exponential distribution to *n*_*v*_, the new constant *ν*_*v*_ = 4.01. Clearly, from Fig. 13 we observe a significant drop in the number of secondary infections and superspreading events. The pdf of the number of secondary infections with the *δ*-variant with partially effective masks and vaccines is much less stretched than the original variant without masks or vaccine. Still the finite risk of superspreading event sustains. However, with 80% vaccination and 50% reduced occupancy, coupled with masks, a significant reduction in overdispersion is attained. This behooves the need for rapidly vaccinating the population alongside physical intervention measures like high-quality masks, reduced occupancy, and across the board higher ventilation rates.

## 4 Conclusions

Overdispersion leading to superspreading events is a major driving force of the Covid-19 pandemic. Understanding the factors that lead to overdispersion in individual infectivity are also considered long standing scientific problems. Coupling an aerosol mixing model with real-world inputs: exhaled aerosol size distribution for speech and breath, measured viral load distribution, occupancy information from SafeGraph for full-service restaurants in major US cities, realistic ventilation rate distributions, we explore the overdispersion in the number of secondary infections per infector. The simulated results demonstrate that aerosol transmission route is consistent with the overdispersed individual infectivity with viral load variability being a dominant factor that controls secondary attack rates alongside ventilation rate, exposure time, and speaking time. We derive analytical expressions that can accurately and probably for the first time, describe the simulated pdfs of the secondary attack rates and number of secondary infections, respectively. The function for the secondary infection number pdf elucidates quantitatively, how overdispersed distributions of viral load and occupancy, jointly control the overdispersion in the number of secondary infections per infector. Finally, given the input data, modeling assumptions and the scope of the study, it appears significant reduction in transmissibility (both the average, as well as the dispersion) of the highly transmissible *δ*−variant necessitates all possible mitigation measures: efficient masks, high ventilation rates, and reduced occupancy, even after a significant fraction of the population has been vaccinated.

## Data Availability

Data not protected by agreements could be made available upon reasonable request to the corresponding author.

## 5 Acknowledgements

UofT researchers acknowledge support from the Fields Institute for Research in Mathematical Sciences through their project Mathematics for Public Health and Variants of Concern sponsored by the Canadian Institutes of Health Research. SC acknowledges the Heuckroth Distinguished Faculty Award in Aerospace Engineering from UTIAS. SM acknowledges the Tier 2 Canada Research Chair in Mathematical Modeling and Program Science (grant No. CRC-950-232643).

