## Supplementary Material for "Analysis of overdispersion in airborne transmission of Covid-19"

#### 1 Particle size distribution

From Pöhlker et al. [1] we have the number size distribution as

$$\psi = \sum_{i=1}^{n} A_i \exp\left( - \left[ \frac{\ln(D_{s,0}/D_i)}{\sigma_i} \right]^2 \right) \tag{1}$$

where they have defined $\psi = d\eta/dlogD_{s,0}$. $D_i$ is the mode mean geometric diameter, $\sigma_i$ is the modal geometric standard deviation and $A_i$ is the number concentration at $D_i$. These constants are mode specific parameters which have prescribed values for each of the $i$th modes (refer to Table VI in Pöhlker et al. [1] for the values of the constants). Note that the function $\psi$ is not normalized. We require the normalized function $\psi_{D,N} = d\eta/dD_{s,0}$ for our purpose.

$$\psi = \frac{d\eta}{dlogD_{s,0}} = D_{s,0}\frac{d\eta}{dD_{s,0}} = \sum_{i=1}^{n} A_i \exp\left( - \left[ \frac{\ln(D_{s,0}/D_i)}{\sigma_i} \right]^2 \right) \tag{2}$$

It is to be noted that $A_i$ is a number concentration parameter which means $\psi$ would correspond to number of particles per unit volume. Hence, to obtain the distribution for total number of particles ejected per unit time, we have to multiply Eqn. 2 by the total volume exhaled per unit time $\dot{V}$. The $\dot{V}$ values are taken as $360Lh^{-1}$ and $700Lh^{-1}$ for breathing and speaking respectively, both of which fall within the typical range for $\dot{V}$ as mentioned in Table I of Pöhlker et al. [1].

$$\psi_{D,\eta}(D_{s,0}) = \dot{V} \sum_{i=1}^{n} \frac{A_i}{D_{s,0}} \exp\left( - \left[ \frac{\ln(D_{s,0}/D_i)}{\sigma_i} \right]^2 \right) \tag{3}$$

The number size distribution can then be converted to the volume size distribution through the relation

$$\psi_{D,V} = \frac{\pi}{6} {D_{s,0}}^3 \psi_{D,\eta}(D_{s,0}) \tag{4}$$

#### 2 Characteristic deposition velocity

To obtain a unique characteristic deposition velocity over a surface for the entire range of particle size, we first require a functional form for the deposition velocity. Lai and Nazaroff [2] developed a model for said deposition velocity on indoor surfaces which took into account the effects of Brownian diffusion, turbulent diffusion and gravitational settling. They provided a formulation for a quantity $\beta$ (wall deposition coefficient) as a function of the particle diameter. This function can then be put through an averaging operation to obtain a value for the average characteristic deposition velocity $w_d$ as

$$\beta_{avg} = \frac{w_d A}{V} = \frac{(v_{dv})_{avg}A_v + (v_{du})_{avg}A_u + (v_{dd})_{avg}A_d}{V} \tag{5}$$

Here $A_v$, $A_u$ and $A_d$ are the surface areas of the vertical surfaces, upward-facing horizontal surfaces and downward-facing horizontal surfaces respectively.The corresponding characteristic deposition velocities are given by $v_{dv}$, $v_{du}$ and $v_{dd}$. Lai and Nazaroff [2] write the wall deposition coefficient $\beta$ as a function of the deposition velocities and the room geometry as

$$\beta = \frac{v_{dv}A_v + v_{du}A_u + v_{dd}A_d}{V} \tag{6}$$

The deposition velocities can be expressed in terms of the friction velocity $u^*$ and gravitational settling velocity $u_s$ as

$$v_{dv} = \frac{u^*}{I}, \quad v_{du} = \frac{v_s}{1 - \exp\left(-\frac{v_s I}{u^*}\right)}, \quad v_{dd} = \frac{v_s}{\exp\left(\frac{v_s I}{u^*}\right) - 1} \tag{7}$$

$I$ is an integrated quantity (refer [2] for detailed formulation) that depends on the Schmidt number $Sc = \nu/D$ where $\nu$ is the kinematic viscosity and $D$ is the Brownian diffusivity. Brownian diffusivity can be directly obtained [3] from the particle diameter at equilibrium $D_s = D_{s,0}/5$, as

$$D = \frac{k_B T C_c}{3\pi\mu D_s} \tag{8}$$

Here $k_B$ is the Boltzmann constant, $T$ is the absolute temperature in Kelvin, $\mu$ is the dynamic viscosity of air and $C_c$ is a slip correction factor for small particles. For the remaining two unknowns, the friction velocity and the gravitational settling velocity, the former is an input variable while the later can be expressed as [3]

$$u_s = \frac{\rho {D_s}^2 g C_c}{18\mu} \tag{9}$$

where $\rho$ is the particle density. From Eqn. 6 - 9 an average wall deposition velocity on $i$th surface (where $i = v, u, d$), $(v_{di})_{avg}$ can be obtained by treating this quantity as the expected value of a function of a random variable $(D_s)$ .

$$(v_{di})_{avg} = \int_{-\infty}^{\infty} v_{di}(D_s)\psi_{D_s}(D_s)dD_s \tag{10}$$

It is to be noted that this is a volume averaging operation due to $\psi_{D_s}$ being defined as a volume size distribution similar to Eqn. 4 but by using the equilibrium diameter $D_s$ instead. This gives us a relation for $\beta_{avg}$ that can in turn be related to the characterisitc deposition velocity $w_d$ as

$$\beta_{avg} = \frac{w_d A}{V} = \frac{(v_{dv})_{avg}A_v + (v_{du})_{avg}A_u + (v_{dd})_{avg}A_d}{V} \tag{11}$$

### 3 Exposure time, speaking time, and ACH

Exposure time $\tau$ variation was modeled with a log-normal distribution such that median $\tau = 1$ hr, $(\mu_\tau = 0, \sigma_\tau = 0.40)$ and speech time $t_s$ was modeled using a uniform distribution. In particular $0.25\tau \le t_s \le 0.5\tau$. These yield the following averages: $\langle \tau \rangle = 1.0875$ hr and $\langle t_s \rangle = 0.409$ hr, respectively. From the literature it is found that ventilation rates in indoor built environments are typically log-normally distributed. In particular Bohanon et al. [4] measured the ventilation rate in 33 restaurants and found them to be log-normally distributed with a median 1.8 l/sm$^2$ and standard deviation of 2.1 l/sm$^2$. Following this measured distribution, for $H = 3$m, we found median$ACH$= 2.16 hr$^{-1}$, and used $\mu_{ACH} = 0.7701, \sigma_{ACH} = 0.7554$ for the$ACH$pdf.

### 4 Input parameter values

Table 1: Input Parameters

| **Symbol** | **Definition** | **Value** | **Reference** |
|---|---|---|---|
| $N_s$ | Total number of random samples used | 103,679 | Total number of data points from restaurants over 10 US cities |
| $\rho$ | Viral load - log-normal distribution with mean $\mu$ and standard deviation $\sigma$ | $\mu = 13.8394$ and $\sigma = 3.6301$ for original variant | Data from Yang et al. [5] |
| $ACH$ | Air change rate (ACH) - Measure of amount of air replaced in a room per unit time per unit volume of the room, log-normal distribution | $\mu_{ACH} = 0.7701, \sigma_{ACH} = 0.7554$ | Indoor ventilation rate is typically log-normally distributed. Parameters obtained from measured $ACH$ distribution in restaurants from Bohanon et al. [4] |
| $N$ | Susceptible population at a given POI from SafeGraph data, exponential distribution | $\nu = 7.707$ | SafeGraph occupancy and area datasets. |
| $\tau$ | Exposure time of a susceptible individual to the ejected aerosols, log-normal distribution | $\mu_\tau = 0, \sigma_\tau = 0.4$ | Distribution chosen such that average exposure time corresponds to the occupancy time over which the seven day averaged SafeGraph data is obtained. |

| Symbol | Definition | Value | Reference |
|---|---|---|---|
| $t_s$ | Duration of speaking of the infected individual, uniform distribution | $0.25\tau \leq t_s \leq 0.5\tau$ | Estimated such that $t_s \leq 0.5\tau$ to ensure speaking time is shared between infectors and susceptibles. |
| $D_{s,0}$ | Initial droplet diameter distribution (lognormal) | $10^{-2}\mu m \leq D_{s,0} \leq 10^{2}\mu m$ | Range used by Pöhlker et al. [1] |
| $D_i$ | Mean geometric diameter of a mode of expiratory event ($b$ - breath, $s$ - speech) | $(D_1)_b = 0.07\mu m$, $(D_2)_b = 0.3\mu m$, $(D_1)_s = 0.07\mu m$, $(D_2)_s = 0.3\mu m$, $(D_3)_s = 1.00\mu m$, $(D_4)_s = 10\mu m$, $(D_5)_s = 96\mu m$ | Input parameter in the lognormal fit function for respiration PSDs provided by Pöhlker et al. [1] (Table VI). |
| $A_i$ | Number concentration at $D_i$ | $(A_1)_b = 7.7cm^{-3}$, $(A_2)_b = 1.1cm^{-3}$, $(A_1)_s = 9.8cm^{-3}$, $(A_2)_s = 1.4cm^{-3}$, $(A_3)_s = 1.7cm^{-3}$, $(A_4)_s = 0.03cm^{-3}$, $(A_5)_s = 0.17cm^{-3}$ | Input parameter in the lognormal fit function for respiration PSDs provided by Pöhlker et al. [1] (Table VI). |
| $\sigma_i$ | Modal geometric standard deviation | $(\sigma_1)_b = 0.9$, $(\sigma_2)_b = 0.9$, $(\sigma_1)_s = 0.9$, $(\sigma_2)_s = 0.9$, $(\sigma_3)_s = 0.9$, $(\sigma_4)_s = 0.98$, $(\sigma_5)_s = 0.97$ | Input parameter in the lognormal fit function for respiration PSDs provided by Pöhlker et al. [1] (Table VI). |
| $\dot{V}$ | Volume of air exhaled in an expiratory event ($b$ - breath, $s$ - speech) | $\dot{V}_s = 194.4cm^3s^{-1}$, $\dot{V}_b = 100cm^3s^{-1}$ | Characteristic values used for calculations in Pöhlker et al. [1] (Table I). |

| Symbol | Definition | Value | Reference |
| --- | --- | --- | --- |
| $\dot{V}_b$ | Inhaled volume of air | $100cm^3s^{-1}$ | Inhalation and exhalation volume during one breath cycle considered to be equal. Value for exhalation volume obtained from Pöhlker et al. [1] (Table I). |
| $H$ | Room height | $3m$ | |
| $u^*$ | Friction velocity | $0.03ms^{-1}$ | Chosen from the range of values used in Lai and Nazaroff [2]. Fig.4 [2] shows that all values in the range has overlapping deposition velocity curves in the higher particle size range, which determines the average deposition velocity value. |
| $T$ | Ambient temperature | $294.7K$ | Recommended typical indoor condition by ASHRAE |
| $RH$ | Relative humidity | 0.5 | Recommended typical indoor condition by ASHRAE |
| $UV_{index}$ | Strength of ultraviolet radiation | 0 | Recommended typical indoor condition by ASHRAE |

| Symbol | Definition | Value | Reference |
|---|---|---|---|
| $t_{1/2}$ | Half-life of airborne virus within aerosols | $32.07 min$ | Based on above parameters at standard indoor conditions, from Dabisch et al. estimated using DHS calculator [6]. [7] |
| $r_v$ | Dose response constant | 1/1440 *copies* | Obtained from Schijven et al. [8] |

### 5 Sensitivity of mean and standard deviation of number of secondary infections

In this sub-section we explore the sensitivity of the most important parameters: $\langle ACH \rangle$, $\langle \rho \rangle$, and $\langle N \rangle$. To that end we explore effect of variation one parameter at a time on $\langle Z \rangle$ and standard deviation $(Z)$, keeping the other two constant. The results are shown in Table 2 and Figs. 1a, 1b, and 1c. All three parameters affect $\langle Z \rangle$ and standard deviation $(Z)$. However, the effect of variation of $\langle \rho \rangle$ is strongest on both the mean and standard deviation of $Z$, followed by that of $\langle N \rangle$, and $\langle ACH \rangle$.

Table 2: Sensitivity of secondary infection number statistics

| $\langle ACH \rangle$ | $\alpha$ | $\mu$ | $\sigma$ | $\langle \rho \rangle$ | $\langle N \rangle$ | $\langle Z \rangle$ | **std**$(Z)$ |
|---|---|---|---|---|---|---|---|
| 0.1 | 1.480x$10^{-10}$ | 13.839 | 3.643 | 7.796x$10^{8}$ | 7.707 | 0.149 | 1.099 |
| 1 | 1.340x$10^{-10}$ | 13.839 | 3.643 | 7.796x$10^{8}$ | 7.707 | 0.141 | 1.061 |
| 5 | 9.470x$10^{-11}$ | 13.839 | 3.643 | 7.796x$10^{8}$ | 7.707 | 0.113 | 0.933 |
| 10 | 6.920x$10^{-11}$ | 13.839 | 3.643 | 7.796x$10^{8}$ | 7.707 | 0.091 | 0.828 |
| 50 | 2.180x$10^{-11}$ | 13.839 | 3.643 | 7.796x$10^{8}$ | 7.707 | 0.041 | 0.519 |
| 100 | 1.170x$10^{-11}$ | 13.839 | 3.643 | 7.796x$10^{8}$ | 7.707 | 0.026 | 0.395 |
| 2 | 1.220x$10^{-10}$ | 12.672 | 3.643 | 2.427x$10^{8}$ | 7.707 | 0.061 | 0.653 |
| 2 | 1.220x$10^{-10}$ | 14.672 | 3.643 | 1.793x$10^{9}$ | 7.707 | 0.221 | 1.370 |

| $\langle ACH \rangle$ | $\alpha$ | $\mu$ | $\sigma$ | $\langle \rho \rangle$ | $\langle N \rangle$ | $\langle Z \rangle$ | $\mathbf{std}(Z)$ |
|---|---|---|---|---|---|---|---|
| 2 | $1.220 \text{x} 10^{-10}$ | 16.672 | 3.643 | $1.325 \text{x} 10^{10}$ | 7.707 | 0.644 | 2.484 |
| 2 | $1.220 \text{x} 10^{-10}$ | 18.672 | 3.643 | $9.789 \text{x} 10^{10}$ | 7.707 | 1.492 | 3.859 |
| 2 | $1.220 \text{x} 10^{-10}$ | 20.672 | 3.643 | $7.234 \text{x} 10^{11}$ | 7.707 | 2.698 | 5.133 |
| 2 | $1.220 \text{x} 10^{-10}$ | 21.672 | 3.643 | $1.966 \text{x} 10^{12}$ | 7.707 | 3.294 | 5.597 |
| 2 | $1.220 \text{x} 10^{-10}$ | 13.839 | 3.643 | $7.796 \text{x} 10^{8}$ | 1.707 | 0.029 | 0.226 |
| 2 | $1.220 \text{x} 10^{-10}$ | 13.839 | 3.643 | $7.796 \text{x} 10^{8}$ | 2.707 | 0.046 | 0.359 |
| 2 | $1.220 \text{x} 10^{-10}$ | 13.839 | 3.643 | $7.796 \text{x} 10^{8}$ | 4.707 | 0.081 | 0.625 |
| 2 | $1.220 \text{x} 10^{-10}$ | 13.839 | 3.643 | $7.796 \text{x} 10^{8}$ | 6.707 | 0.115 | 0.891 |
| 2 | $1.220 \text{x} 10^{-10}$ | 13.839 | 3.643 | $7.796 \text{x} 10^{8}$ | 8.707 | 0.149 | 1.156 |
| 2 | $1.220 \text{x} 10^{-10}$ | 13.839 | 3.643 | $7.796 \text{x} 10^{8}$ | 10.707 | 0.184 | 1.422 |

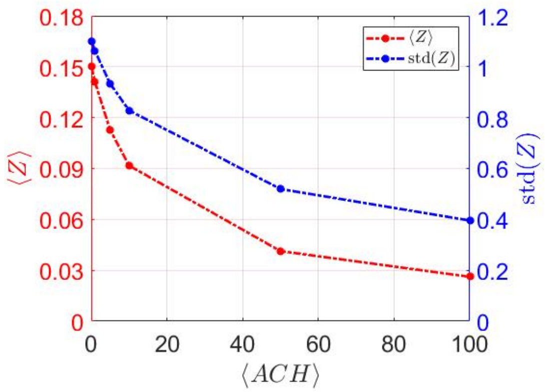


(a) $ACH$ sensitivity

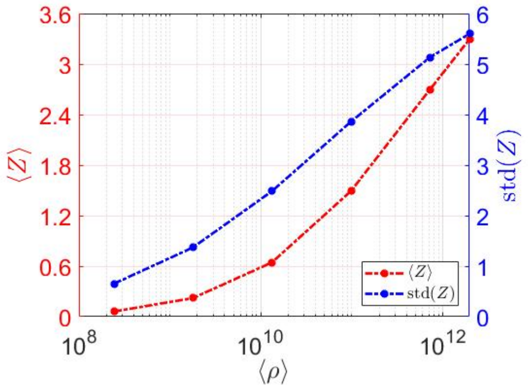


(b) $\rho$ sensitivity

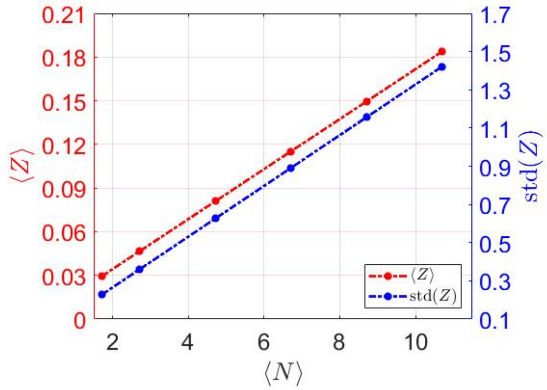


(c) $N$ sensitivity

Figure 1: Variation of mean $\langle Z \rangle$ and standard deviation $std(Z)$ with (a)mean air-change rate, (b)mean viral load and (c)mean occupancy.

### 6 Effect of masking, fixed ventilation rates, vaccines, and reduced occupancy - inputs and results

Table 3: Effect of masking, fixed ventilation rates, vaccines, and reduced occupancy - inputs and results. Note that $f_{mask}$ corresponds to the percentage of aerosol concentration a mask is able to block and $f_{occ}$ refers to the percentage of original occupancy.

| **Parameter** | **Original variant, no masks** | **$\delta$–variant, no masks** | **Original variant, ACH** $= 5$, **mask** 50% | **$\delta$–variant, ACH** $= 5$, **mask** 50% | **$\delta$–variant, Vaccine,** 100% **occupancy** | **$\delta$–variant, Vaccine,** 50% **occupancy** |
|---|---|---|---|---|---|---|
| $\mu$ | 13.839 | 20.672 | 13.839 | 20.672 | 20.672 | 20.672 |
| $\sigma$ | 3.642 | 3.642 | 3.642 | 3.642 | 3.642 | 3.642 |
| $\eta_{vac}$ | 0.6 | 0.6 | 0.6 | 0.6 | 0.6 | 0.6 |
| $\eta_{cov}$ | 0 | 0 | 0 | 0 | 0.8 | 0.8 |
| $F_{occupancy}$ | 1 | 1 | 1 | 1 | 1 | 0.5 |
| $F_{mask}$ | 1 | 1 | 0.5 | 0.5 | 0.5 | 0.5 |
| $\langle Z \rangle$ | 0.125 | 2.648 | 0.043 | 1.693 | 0.931 | 0.465 |
| **std**$(Z)$ | 0.994 | 5.090 | 0.537 | 4.112 | 2.198 | 1.099 |
| $\mathcal{Z}$, CI 5% | 8.543x$10^{-7}$ | 7.954x$10^{-4}$ | 1.876x$10^{-7}$ | 1.692x$10^{-4}$ | 1.043x$10^{-4}$ | 5.242x$10^{-5}$ |
| $\mathcal{Z}$, CI 95% | 0.278 | 12.923 | 0.058 | 9.361 | 5.072 | 2.549 |
| $\mathcal{Z}$, CI 99% | 2.963 | 24.366 | 0.748 | 20.516 | 10.881 | 5.469 |
